# Patterns of Dysglycemia Identified by Continuous Glucose Monitoring among Critically Ill Children in Malawi and Bangladesh

**DOI:** 10.64898/2026.01.30.26344670

**Authors:** Philliness P. Harawa, Celine Bourdon, Farnaz Khoshnevisan, Shafiqul A. Sarker, Munirul Islam, Farhana Islam, Zahidul Islam, Chikondi Makwinja, Emmanuel Chimwezi, Narshion Ngao, Caroline Tigoi, Shaima S. Nahar, James Chirombo, Guanlan Hu, Paraskevi Massara, Stanley Khoswe, Emmie Mbale, Edward Senga, Benjamin Kumwenda, Tahmeed Ahmed, Judd L. Walson, James A. Berkley, Mohammed J. Chisti, Wieger P. Voskuijl, Farzana Afroze, Robert H.J. Bandsma

**Affiliations:** The Childhood Acute Illness and Nutrition (CHAIN) Network, Nairobi, Kenya; Department of Paediatrics and Child Health, Kamuzu University of Health Sciences, Blantyre, Malawi; Translational Medicine Program, Hospital for Sick Children, Toronto, Canada; Department of Nutritional Sciences, Faculty of Medicine, University of Toronto, Toronto, Ontario, Canada; Nutrition Research Division (NRD), International Centre for Diarrhoeal Disease Research, Bangladesh (icddr,b), Dhaka, Bangladesh; Department of Biomedical Sciences, Kamuzu University of Health Sciences, Blantyre, Malawi; Clinical Research Department, KEMRI–Wellcome Trust Research Programme, Kilifi, Kenya; Centre for Tropical Medicine and Global Health, University of Oxford, Oxford, United Kingdom; Malawi-Liverpool Wellcome Trust Programme, Blantyre, Malawi; Departments of International Health, Medicine, Pediatrics, Johns Hopkins University; Department of Paediatrics, Amsterdam Centre for Global Child Health, Emma Children’s Hospital, Amsterdam UMC, University of Amsterdam, Amsterdam, Netherlands; Division of Paediatric Gastroenterology, Hepatology and Nutrition, The Hospital for Sick Children, Toronto, Canada; Centre for Global Child Health, Hospital for Sick Children, Toronto, Canada

**Keywords:** Glucose, “Continuous glucose monitoring”, Hypoglycemia, Hyperglycemia, “Glucose variability”, Wasting, Kwashiorkor, “Edematous malnutrition”, Malnutrition, Children, “Acute illness”, “Critical illness, Hospital, Africa, “South Asia”, severe acute malnutrition

## Abstract

Dysglycemia is a critical metabolic disturbance associated with mortality in acutely ill children, yet its burden may be underrecognized in low-income settings due to reliance on single point-of-care measurements. Using continuous glucose monitoring (CGM), we aimed to characterize glucose patterns in acutely ill children of different anthropometric status.

**Methods:** Children aged 2-23 months admitted with acute illness were prospectively recruited from two hospitals in Bangladesh and Malawi. Clinical data were collected, and interstitial glucose was monitored for 48 hours using the Dexcom G4 Platinum system. Glucose excursions and variability were analyzed and associated with anthropometric status.

**Results:** Of 93 enrolled children, 88 had sufficient CGM data: 21 not wasted (NW), 22 moderately wasted (MW), and 45 with severe malnutrition (SAM; 29 severe wasting [SW], 16 edematous malnutrition [EM]). Low-glucose excursions were detected in 8 (38%) children with NW, 11 (50%) with MW, 12 (41%) with SW, and 10 (63%) with EM. While not confirmed hypoglycemia, these low-glucose excursions were longer and more frequently below severe thresholds in children with EM. Hyperglycemic excursions occurred in 31% of children and were longer in children with SAM compared to NW (median 41 vs. 23 min, p<0.0001). Overall, 35% of children maintained euglycemic profiles, while others exhibited marked glucose variability.

**Conclusion:** CGM revealed frequent glucose instability among acutely ill children, with patterns varying across anthropometric groups. When interpreted cautiously, CGM may serve as a research tool to detect dysglycemia and assess response to therapeutic or nutritional interventions in critically ill children in low-resource settings.

## Background

The first 48 hours of hospital admission represent a critical period for acutely ill children, during which life-threatening complications often arise from the interplay of infection, inflammation, and metabolic instability^1–7^ Dysglycemia is a serious metabolic disturbance commonly observed in critically ill children, with both hypo- and hyperglycemia associated with adverse clinical outcomes.^8–12^ Mortality risk follows a J- or U-shaped relationship with glycemia.^13–19^ and children with malnutrition are particularly vulnerable to metabolic derangements.^1,17,19–23^ The World Health Organization (WHO) considers all hospitalized children with severe malnutrition to be at risk of hypoglycemia, warranting immediate treatment to prevent death.^24^

Despite recognizing the clinical importance of dysglycemia, data describing its occurrence in acutely ill children in low-resource settings remains limited and heterogeneous. Reported prevalence estimates of hypoglycemia and hyperglycemia in critically ill children vary widely, ranging from 10–25% and 30–80%, respectively.^25^ Among children with severe malnutrition, a meta-analysis estimated the prevalence of hypoglycemia to be 9%^26^ but studies report a range from 4-14%.^16,27–30^ Higher estimates have been reported when less stringent glycemic thresholds were applied; for example, a Nigerian study reported a prevalence of 21% using a threshold of <3.6 mmol/L.^31^ Hypoglycemia in acutely ill children with severe malnutrition was associated with high case fatality rates, 20 to 44% in studies from Kenya, Tanzania and Nigeria.^4,26–28,31–35^

However, most existing studies have relied on single point-of-care (POC) glucose measurements obtained at admission or during overt clinical deterioration suggestive of hypoglycemia. This symptom-based screening approaches is limited, as clinical signs such as lethargy or seizures are non-specific and may be transient or absent. Consequently, clinically relevant dysglycemia may go undetected, particularly among high-risk children with sepsis and/or low anthropometric status, and opportunities for intervention can be missed. As a result, the temporal patterns and burden of dysglycemia, as defined by the WHO, among critically ill children in low-resource settings remains largely unknown, particularly among those with moderate or no malnutrition.

Continuous glucose monitoring (CGM), originally developed for diabetes management, enables high-frequency assessment of interstitial glucose and provides information on temporal glucose patterns. In high-resource settings, CGM is increasingly used to identify dysglycemic patterns in critically ill children.^36–38^ However, to date, CGM has not been systematically applied to hospitalized children in low-resource settings, a vulnerable population with a broader range of anthropometric statuses.

In this study, we analyzed 48 hours of CGM data captured early during hospital admission to characterize glucose patterns, including variability and low- or high-glucose excursions among acutely ill children across anthropometric statuses in Bangladesh and Malawi. By examining glucose instability in relation to anthropometric status, this work provides insight into dysglycemic patterns within a high-risk pediatric population. Although CGM has methodological limitations for estimating the true incidence of hypoglycemia, this work may inform future studies that use CGM to characterize dynamic glucose responses to therapeutic or nutritional interventions for critically ill children in low-resource settings.

## Methods

### Study setting and population

This observational prospective cohort study was conducted within the framework of the Childhood Acute Illness and Nutrition (CHAIN) cohort.^3,39^ The CGM sub-study was designed to assess glucose homeostasis across a range of anthropometric statuses in acutely ill children. Between September 2018 and April 2020, we recruited children aged 2-23 months from two sites: Queen Elizabeth Central Hospital (QECH), Blantyre, Malawi and the International Centre for Diarrhoeal Disease Research, Bangladesh (icddr,b) Hospital, Dhaka, Bangladesh. QECH is the largest tertiary hospital in Malawi with approximately 26,000 pediatric admission per year, including 1,200 children with severe malnutrition.^40,41^ The icddr,b Hospital specializes in diarrhoeal disease and provides care to over 120,000 children each year, of whom approximately 50% present with severe malnutrition.^42^ Supplemental Materials detail the exclusion and inclusion criteria (Section 2) and the recruitment process (Section 1) which were as per CHAIN protocol.^39^ Enrolment into this CGM sub-study was convenience-based.

Ethical approval was granted from: 1) Oxford University Tropical Research Ethics Committee, U.K.; 2) the Research Ethics Board of the Hospital for Sick Children, Canada; 3) icddr,b Research Review Committee and Ethical Review Committee, Bangladesh; 4) the QECH hospital director and Paediatric department, Malawi; and 5) College of Medicine Research and Ethics Committee Kamuzu University of Health Sciences, Malawi. Written informed consent was obtained from each child’s legal guardian prior to enrollment, either by signature or witnessed thumbprint. Consent forms were translated into local languages to ensure comprehension. For caregivers unable to read, study information was read aloud and explained in the presence of an impartial witness.

### Anthropometry

Anthropometric status was evaluated at admission. Children were classified into four anthropometric groups: no wasting (NW), moderate wasting (MW), severe wasting (SW), or nutritional edema (EM) (**Supplementary Table 1**). Measures of mid upper arm circumference (MUAC), weight, and length were taken using non-stretch tape (TALC, St. Albans, UK), length boards (Secca 416 infantometer, Birmingham, UK), and digital weight scales (Seca 825, calibrated monthly). MUAC and length were taken by two independent observers and repeated if discrepant by >5 mm or >7 mm, respectively.

### Clinical procedures and data collection

Before study initiation, all procedures and clinical definitions were harmonized across sites through cross-site training, adherence to standard operating procedures, and regular internal and external audits.^43^ Identical equipment was procured and used. Clinicians and research staff collected data on standardized case report forms, available online at https://chainnetwork.org/resources/. Comprehensive clinical data was collected including initial presentation, socio-demographics, clinical history, blood biochemistry and complete blood count, HIV and malaria status. Clinical progression was documented daily. All children received care according to current national and international clinical guidelines.

### Feeding protocols and standard clinical care

In-hospital feeding protocols followed WHO guidelines for the management of severe malnutrition, with site-specific adaptations during the initial stabilization phase. In Malawi, children with severe malnutrition were fed every three hours using a calculated volume of F75, a commercial milk-based therapeutic formula providing a target energy intake of 80-100 kcal/kg/day.^44^ In F75, approximately 63% of the total energy is derived from carbohydrates. Moderately wasted and non-malnourished children received diets as provided by their family. In contrast, at icddr,b in Bangladesh, all hospitalized children received two-hourly feeds of *milk suji*, a locally prepared alternative to F75. Although considered nutritionally comparable to F75, *milk suji* differs in composition, with carbohydrates accounting for approximately 44% of total energy. In both settings, feeds were given by caregivers under the supervision of nursing staff. In accordance with WHO guidelines, children with severe malnutrition were initiated on broad spectrum antibiotics. Oral rehydration solutions, including ReSoMal, a formulation designed for children with severe malnutrition that contain carbohydrates, were administered to children with diarrhea and/or clinical signs of dehydration.

### Glucose monitoring

POC glucometers (OneTouch, LifeScan IP Holdings, Malvern, PA, USA) were used to measure capillary blood glucose from finger-prick samples obtained at admission, for CGM calibration and to assess hypoglycemic excursions. Within six hours of admission, CGM was started using a pediatric Dexcom G4 platinum system (Dexcom, San Diego, USA). A physician positioned the subcutaneous sensor on the child’s abdomen to measure interstitial glucose at five-minute intervals. CGM calibrations were repeated at least every 12 hours. CGM alerts indicating hypoglycemia were investigated by research staff and verified using with POC glucometers. When hypoglycemia (<3 mmol/L) was confirmed, children were provided with an oral glucose bolus and continued feeds as per standard protocols. Additionally, clinical staff conducted systematic assessments for clinical deterioration, including altered level of consciousness, convulsions, dehydration, respiratory distress and signs of shock (e.g., cool peripheries, weak rapid pulse, capillary refill time >3 seconds).

### Data processing and quality control

Data were entered daily into a centrally hosted REDCap database with built-in internal validity checks. Custom dashboards were developed to enable real-time identification of data irregularities. Variable definitions, data processing steps are detailed in **Supplementary Table 2**. Anthropometric z-scores were derived using the *anthro* R package, based on the 2006 WHO growth standards.^45,46^ Descriptive statistics are presented stratified by anthropometric groups and are reported as counts with percentage or median with interquartile ranges (IQRs). CGM readings beyond device detection range are automatically classified as ‘low’ (<2.2 mmol/L) or ‘high’ (>22.2 mmol/L); these were imputed as 2.0 mmol/l and 22.4 mmol/L, respectively. CGM profiles were excluded if they recorded less than 24 hours of monitoring data, or had poor coverage defined as having less than 60% of expected data during active monitoring. Given the known lag between interstitial glucose (measured by CGM) and capillary blood glucose (measured by POC),^47^ a Bland-Altmann analysis was performed to assess bias between calibration values obtained with POC glucometers and the average of 3 sequential CGM readings spanning a 15 min window.

### Classification of CGM-detected glucose excursions and glycemic variability

Thresholds used to define hypoglycemia vary across studies; however, the WHO definition is a blood glucose level <2.5 mmol/L in non-wasted children and <3.0 mmol/L in those with severe malnutrition.^48^ To facilitate cross-group comparisons, we applied uniform threshold across all participants to classify CGM-detected low glucose values, categorized as: severely low, <2.5 mmol/L; low, <3 mmol/L, and mildly low, <3.9 mmol/L.^49^ Similarly, CGM-detected high glucose were classified as: high, >11 mmol/L and mildly high, >7 mmol/L, consistent with prior literature.^50^ These thresholds were used for analytical comparability and pattern characterization rather than to infer clinical diagnoses.

Indices of glucose variability were derived using the *iglu* R package^51^, including median, interquartile range (IQR), standard deviation (SD), coefficient of variation (CV), average daily risk range (ADRR, i.e., the average of daily extremes (highest and lowest glucose for each day, averaged across days), continuous overall/overlapping net glycemic action (CONGA, i.e., the SD of the difference measured one hour apart, representing short term intra-day variability) and the mean of daily differences (MODD) which captures the mean difference between glucose obtained at the same time on consecutive days, reflecting inter-day variability. As standardized pediatric thresholds for glycemic variability are still evolving, results were contextualized using published values from non-diabetic post-operative children under 12 months of age,^52^ from children under 6 years newly diagnosed with diabetes^53^ or from healthy pediatric populations between 2-15 years.^54–56^ Definitions of all glycemic indices and information regarding normative or comparative values are presented in **Supplemental Table 3**.

### Statistical analysis

Analyses were conducted using a standardized 48-hour CGM recording window beginning at 18:00 on the first day of admission to align the diurnal cycle.^57^ This allows assessment of day-night patterns with daytime defined as 06:00 – 22:00. Glucose excursions were evaluated using hurdle models to account for excess zero values, these fit a two-part process to assess: 1) whether the proportion of children experiencing at least one low or high excursion differed between anthropometric groups (binary outcome fit with a binomial distribution), and 2) among children with at least one excursion, whether the number or duration differed (count outcomes modeled using Poisson or negative binomial distributions, as appropriate). Marginal means were derived using *emmeans* R package, with pairwise contracts tested and *p*-values adjusted for multiple comparisons using the Tukey method. Site, sex, and age were considered as covariates in initial models but retained only if contributed meaningfully to model fit. Generalized linear models were fit to compare five metrics of glycemic variability (i.e., CV, SD, ADRR, CONGA and MODD) between anthropometric groups. These analyses were conducted with normalization by active monitoring time to account for variability in the duration of CGM recordings. All statistical analyses and visualizations were performed using R (version 4.0.2).

## Results

Between September 2018 and April 2020, 1690 children were screened at hospital admission, of whom 261 (15%) met eligibility criteria, and caregivers of 105 children provided consent. The study flow diagram is presented in **Figure 1**. CGM data were obtained from 93 patients, but five were excluded because of insufficient monitoring (<24 hours due to early death [n=1] or early discharges [n=2]); or poor monitoring coverage (<60% of expected readings, n=2). Eighty-eight patients with sufficient data (Bangladesh, n=52, Bangladesh; Malawi, n=36) were included in the analysis and classified as NW, n=21; MW, n=22; SW, n=29 and EM, n=16. Baseline characteristics by anthropometric group are presented in **Table 1**. Most children (68%) were younger than 12 months of age, and 75% were breastfeeding at admission. Children with EM were older by 5.6 months (95% CI: 2.6, 8.6) and were largely from Malawi.

**Figure 1.**
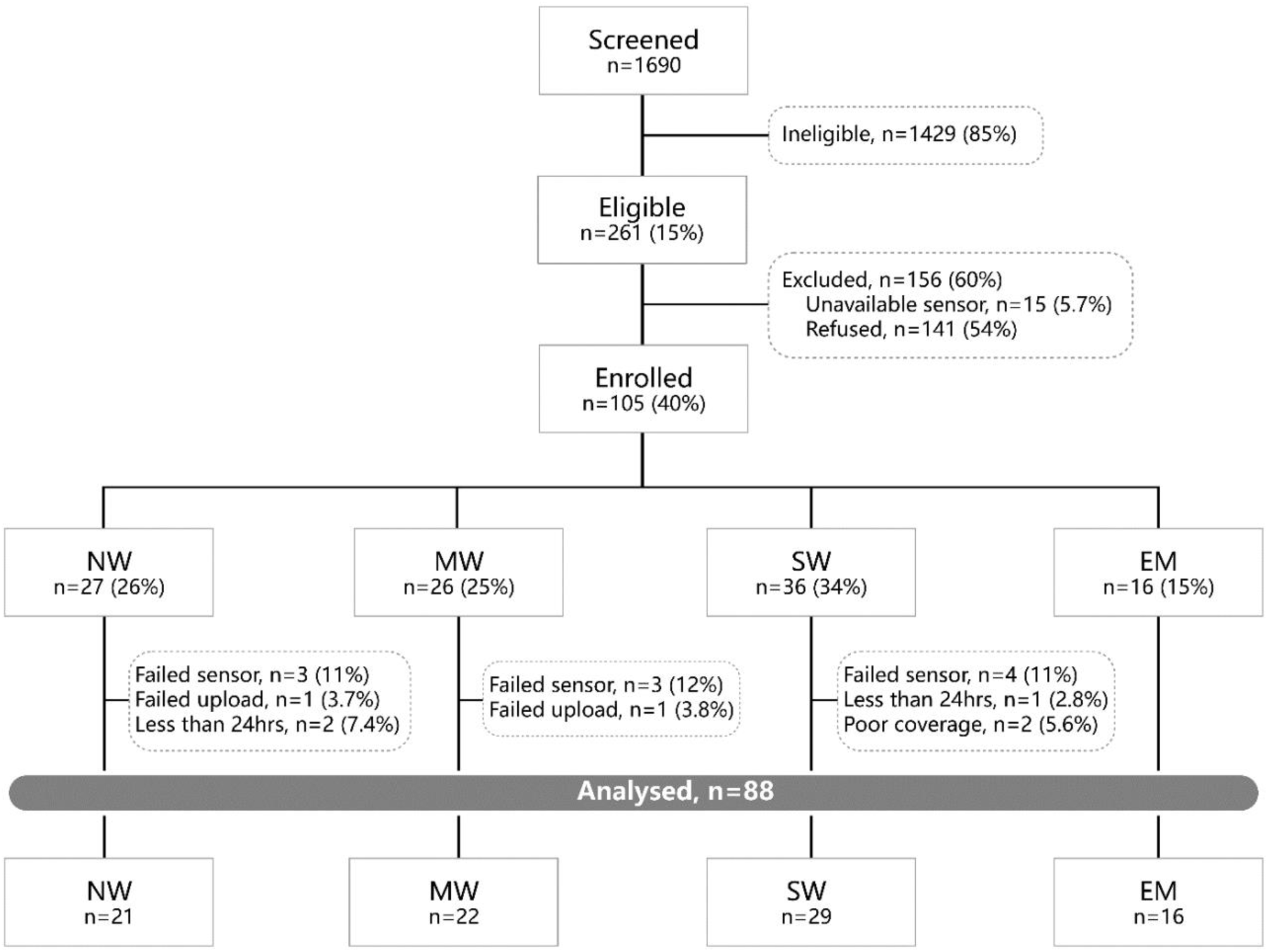
Study flow chart. Children admitted to hospital between September 2018 and April 2020 were screened (n=1690) of which 261 children were eligible. Fifteen participants were not enrolled because sensors were not available due to active usage or machine breakdown. Sensor failure occurred in 10 cases (9.5%). Data upload failed for two participants and was not retrieved. NW, no wasting; MW, moderate wasting; SW, severe wasting; EM, edematous malnutrition.

**Table 1:**
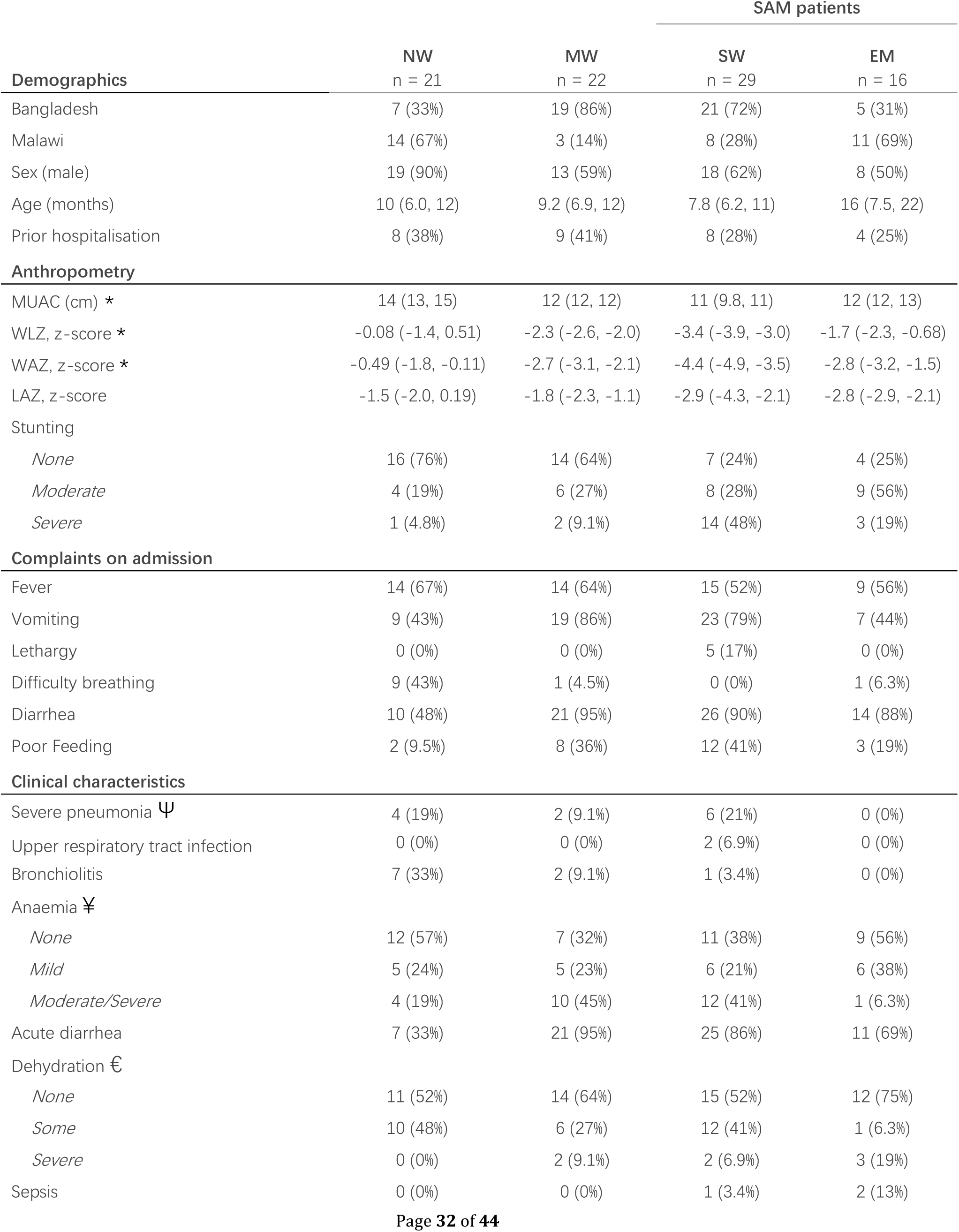
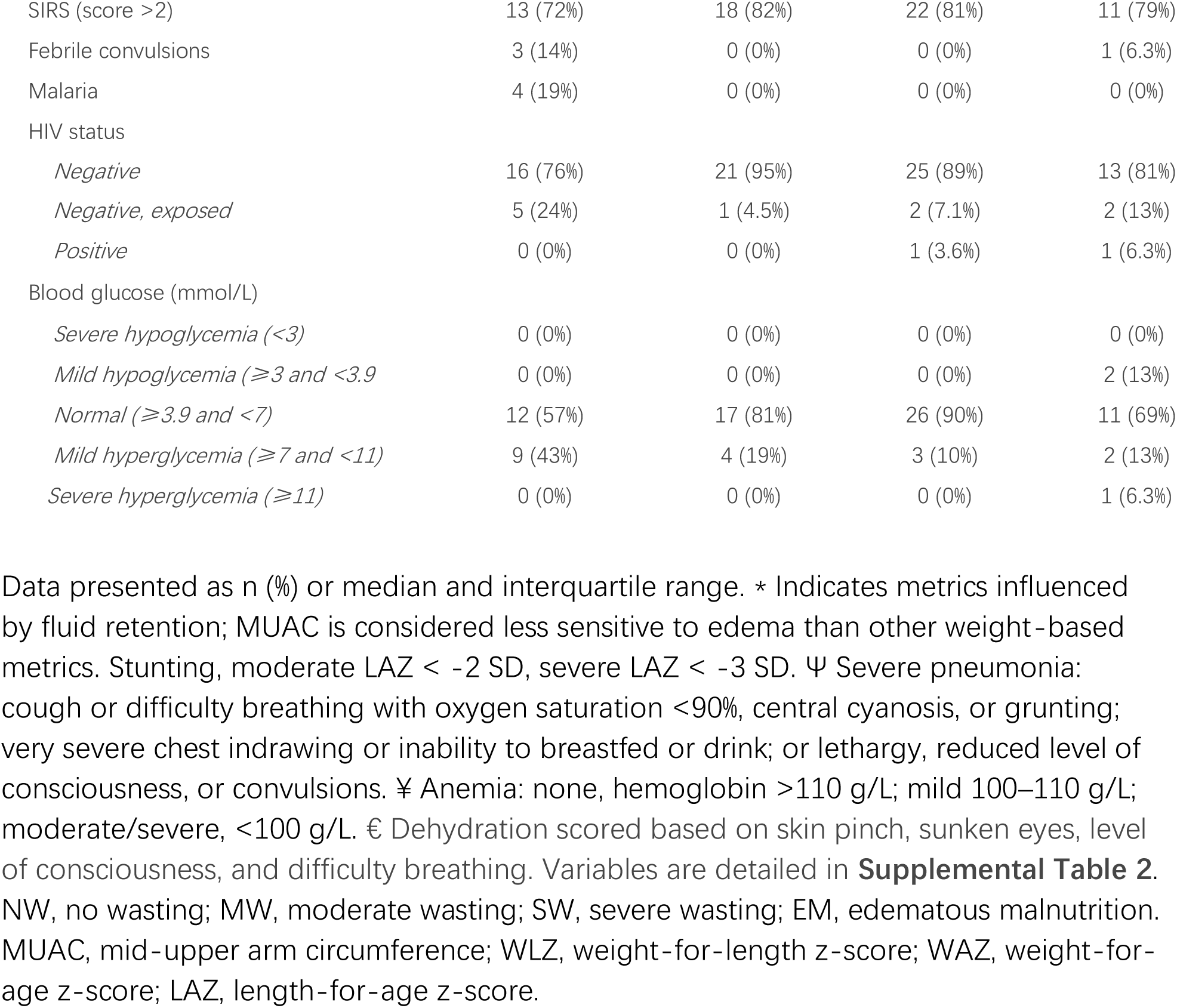
Characteristics of patients at admission stratified by anthropometric groups.

The most common admission diagnoses were gastroenteritis (73%), moderate or severe anemia (42%), and lower respiratory tract infection (27%), with half of patients presenting with two or more of these conditions. Based on POC-measured glucose at admission, dysglycemia was infrequently detected with 2 children (2.3%) classified as having mild hypoglycemia (< 3.9 mmol/L) and 19 (22%) with mild hyperglycemia (≥7 mmol/L).

At admission, children received a range of clinical interventions that could influence glucose homeostasis. A summary of management strategies and clinical outcomes is provided in **Table 2**. Notably, most children at icddr,b (66%) received oral rehydration solutions at admission because of diarrhea severity, while six children (6.8%) received intravenous fluid boluses. Two children (2.3%) received blood transfusions. Length of hospital stay varied by anthropometric status, and three children died during hospitalization.

**Table 2:**
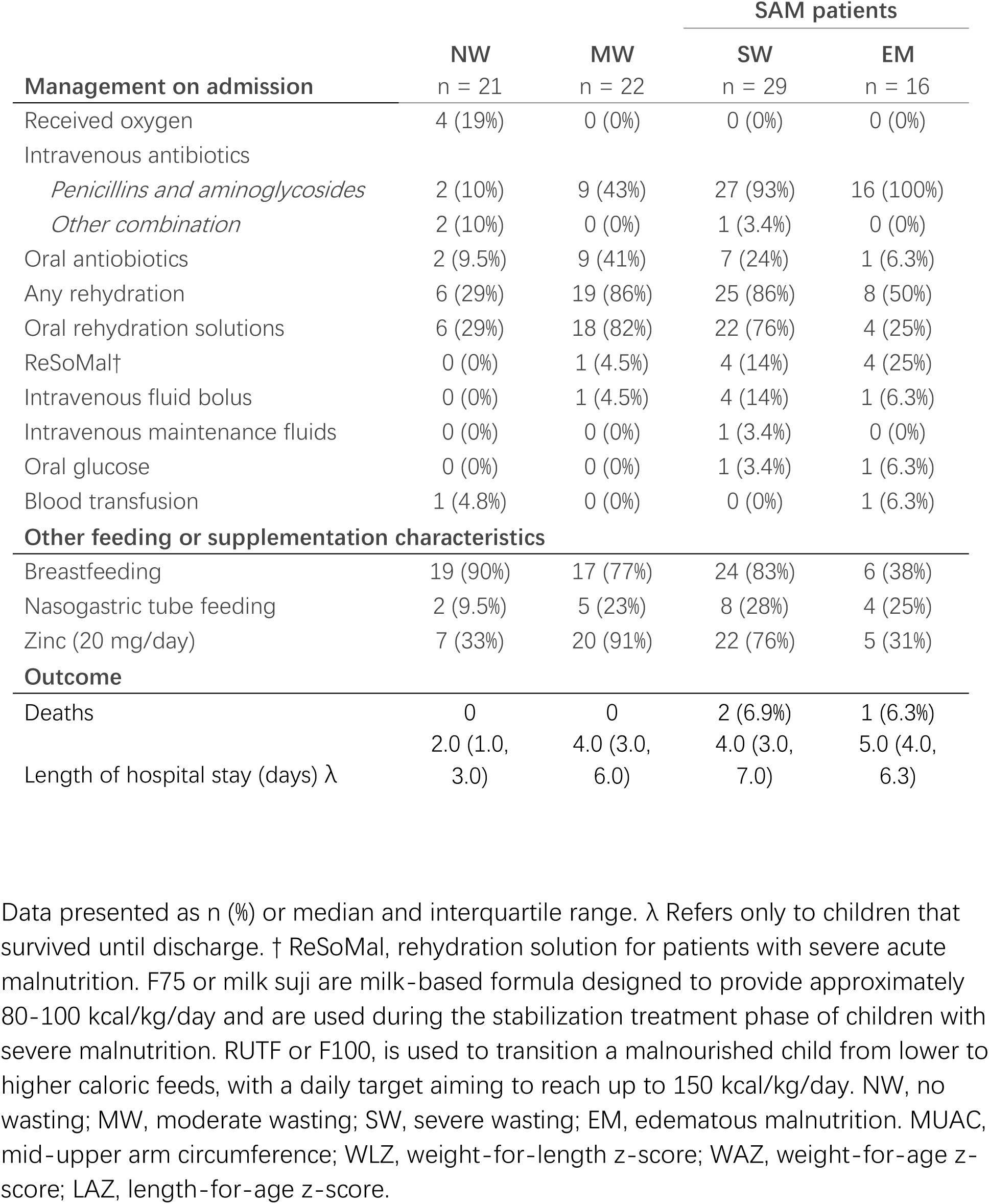
Management and feeding received at admission and outcome of patients stratified by anthropometric groups.

No bias in duration or the proportion of active monitoring was found between anthropometric groups, age groups, or sites.

### Performance of CGM in critically ill children across anthropometric groups

Over the 48-hour CGM recording window, coverage per participant was 97% (95% CI: 96 – 98) % (**Supplemental Figure 1)** which did not differ by anthropometric group, age groups or site. In total, 47,589 CGM readings were obtained (19,526 from 36 children in Malawi and 28,063 from 52 children in Bangladesh). Of these, 874 (1.8%) fell below 3.0 mmol/L, while 1,140 (2.4%) were higher than 11.0 mmol/L (**Figure 1A**). These are not events confirmed by capillary blood measurements and are considered CGM-detected glucose excursions.

To assess potential measurement bias across anthropometric groups, agreement between CGM values and POC measurements was evaluated (n=375 paired observations, Bland-Altman plots presented in **Supplemental Figure 2)**. The limits of agreement spanned approximately ±1.3–1.6 mmol/L but did not differ between groups, while the mean bias between CGM and capillary measurements was small (0.077 mmol/L).

### CGM-detected glucose excursions common in hospitalized children with acute illness

CGM data indicated that many children exhibited glucose patterns consistent with glycemic instability during the 48-hour monitoring window (**Figure 2B-G**). Using hurdle models, we assessed group differences in the probability of experiencing at least one low-or high-glucose excursion and, among affected children, whether the daily number or duration of excursions differed between groups. These analyses were normalized by active monitoring time.

**Figure 2:**
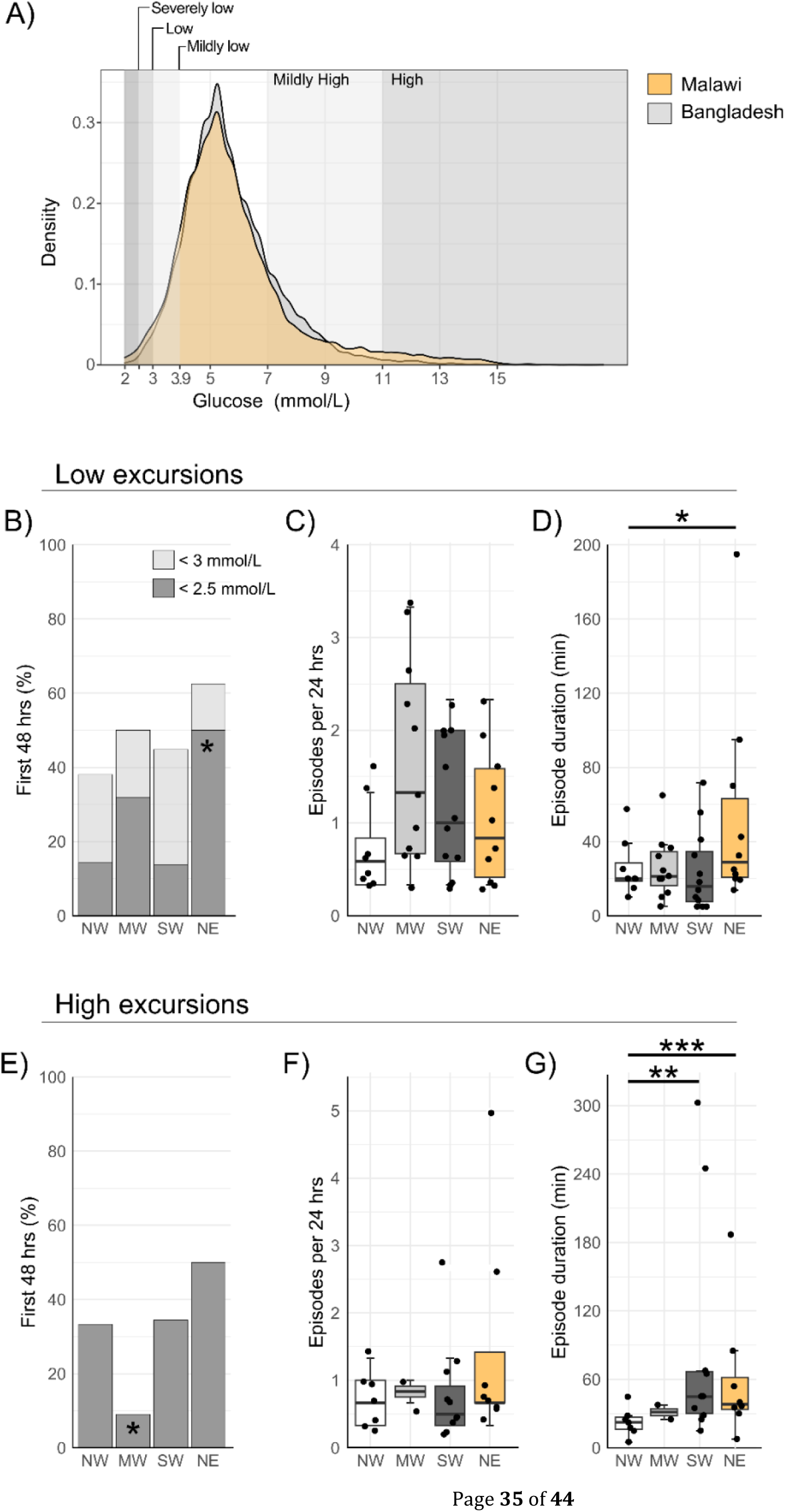
Summary of CGM-detected glucose excursions in the 48 hour monitoring window in severely ill children by anthropometric status. **A**) Density plots of the CGM-measured glucose levels in children from Malawi or Bangladesh. **B-G**) Summary of CGM-detected low and high glucose excursions by anthropometric group. **B)** Proportion of children that experience at least one low excursion (< 3 mmol/L, light grey) or severely low excursion (< 2.5 mmol/L, dark grey); * indicates higher proportion of severely low glucose excursion in children with EM compared to SW, p <0.05. **E**) Proportion of children that experience at least one high excursion (> 11 mmol/L); *indicates a lower proportion in children with MW compared to EM, p <0.05. Boxplots present the median number of daily excursions per participant for **C**) low excursions or **F**) high excursions. Duration of excursions is presented for **D**) low or **G**) high excursions; outliers (> 180 min) were excluded from the analysis to prevent undue influence on duration estimates. Group differences were tested using hurdle models. Significance threshold, *p<0.05, **p<0.01, ***p<0.001. NW, no-wasting; MW, moderate wasting; SW, severe wasting; EM, edematous malnutrition.

Early in admission, 42 children (48%) experienced at least one CGM-detected low glucose excursion (<3.0 mmol/L); with no differences in proportions across groups (**Table 3** and **Supplemental Table 4**). Among children with at least one low excursion, the median number of daily excursions was 0.84 (IQR: 0.37, 2.0). While the frequency of daily excursions did not differ between groups, their duration varied: 24 minutes (95% CI: 20, 27) in NW children, 26 minutes (95% CI: 23, 29) in MW, 24 minutes (95% CI: 21, 27) in SW, and 38 minutes (95% CI: 34, 42) in children with EM. Low glucose excursions were on average 13 minutes longer (95% CI: 8.6, 18) in children with EM compared to other groups. To prevent undue influence, data from one child with EM exhibiting a prolonged low glucose excursion (>180 minutes) was excluded from the duration analysis. Children with EM also tended to experience more excursions crossing the severely low threshold (<2.5 mmol/L), though estimates were imprecise (**Supplemental Table 5)**.

**Table 3.**
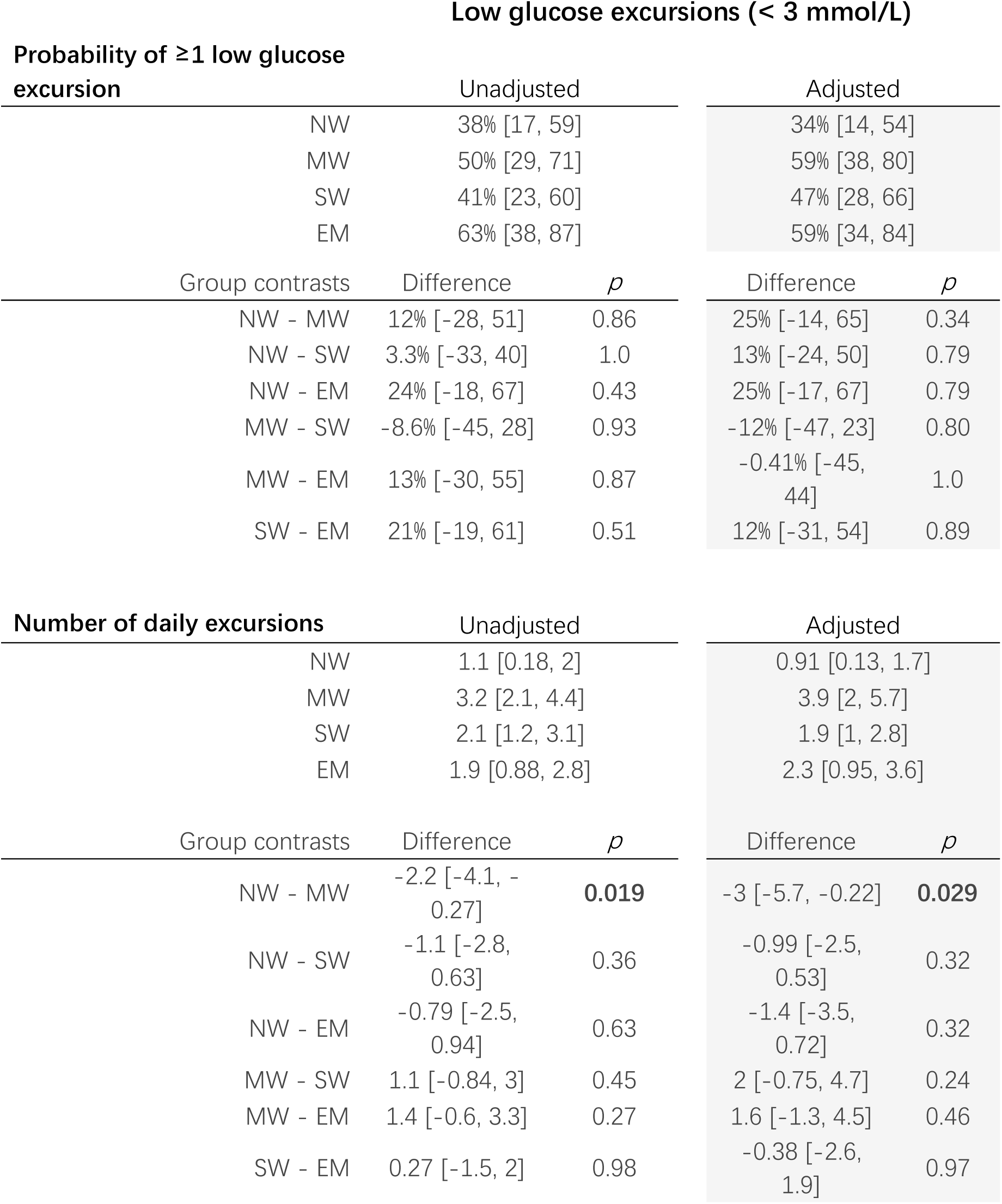
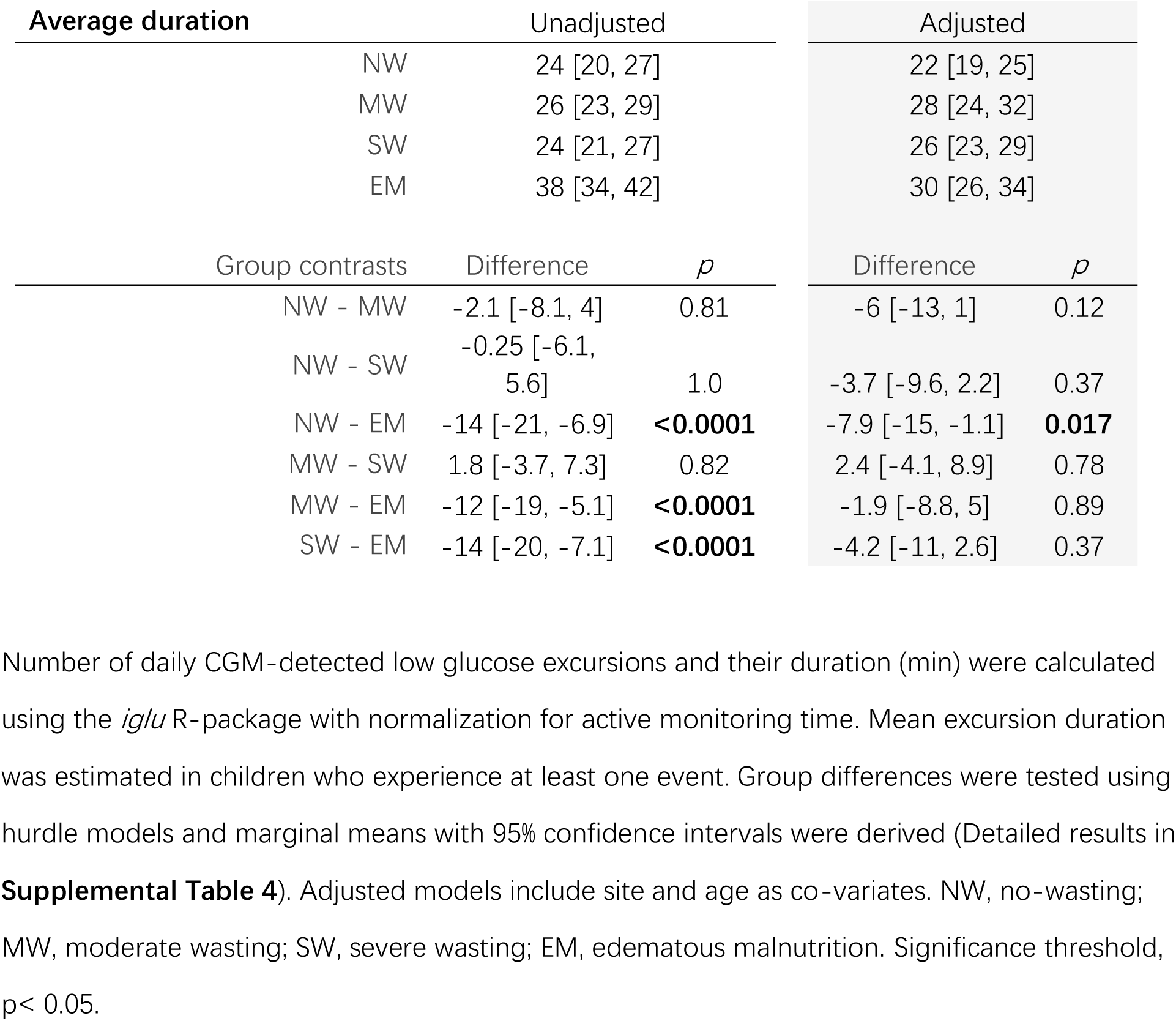
Differences in the probability, frequency, and duration of CGM-detected low glucose excursions (<3.0 mmol/L) during the 48 hour monitoring window by anthropometric status.

After adjustment for age, anthropometric status, and active monitoring time, children in Malawi tended to have a higher probability of experiencing at least one CGM-detected low glucose excursion than those in Bangladesh (62% [95% CI: 45, 80] vs. 37% [95% CI: 22, 52]). Among affected children, daily excursion frequency did not differ by site, but duration was longer in Malawi by a mean of 7.7 minutes (30 [95% CI: 27, 34] vs. 22 [95% CI: 20, 25] min, p = 0.0028).

Early during admission, 27 children (31%) experienced at least one CGM-detected high glucose excursion (>11.0 mmol/L). Proportion varied across anthropometric groups but with overlapping confidence intervals (NW, 33% [95% CI: 13, 54]; MW, 9.1% [95% CI: 0, 21]; SW, 34% [95% CI: 17, 52] and EM, 50% [95% CI: 25, 75], **Table 4; Supplemental Table 6**). Children with MW showed a lower probability of experiencing at least one high excursion compared to children with EM. Among children with at least one excursion, the daily number of high excursions did not differ between groups, but their duration was longer in children with SW (41 min [36, 45]) or EM (41 min [36, 46]) compared to children with NW (23 min [19, 26], both p<0.0001). This analysis excluded 3 children with unusually long excursions (>180 minutes). Overall, high glucose excursions in children with SW or EM lasted approximately 20 minutes longer than those observed in NW children. Full results, including stratification by site and day-night period are provided in **Supplemental Table 6** and **Supplemental Table 7**.

**Table 4.**
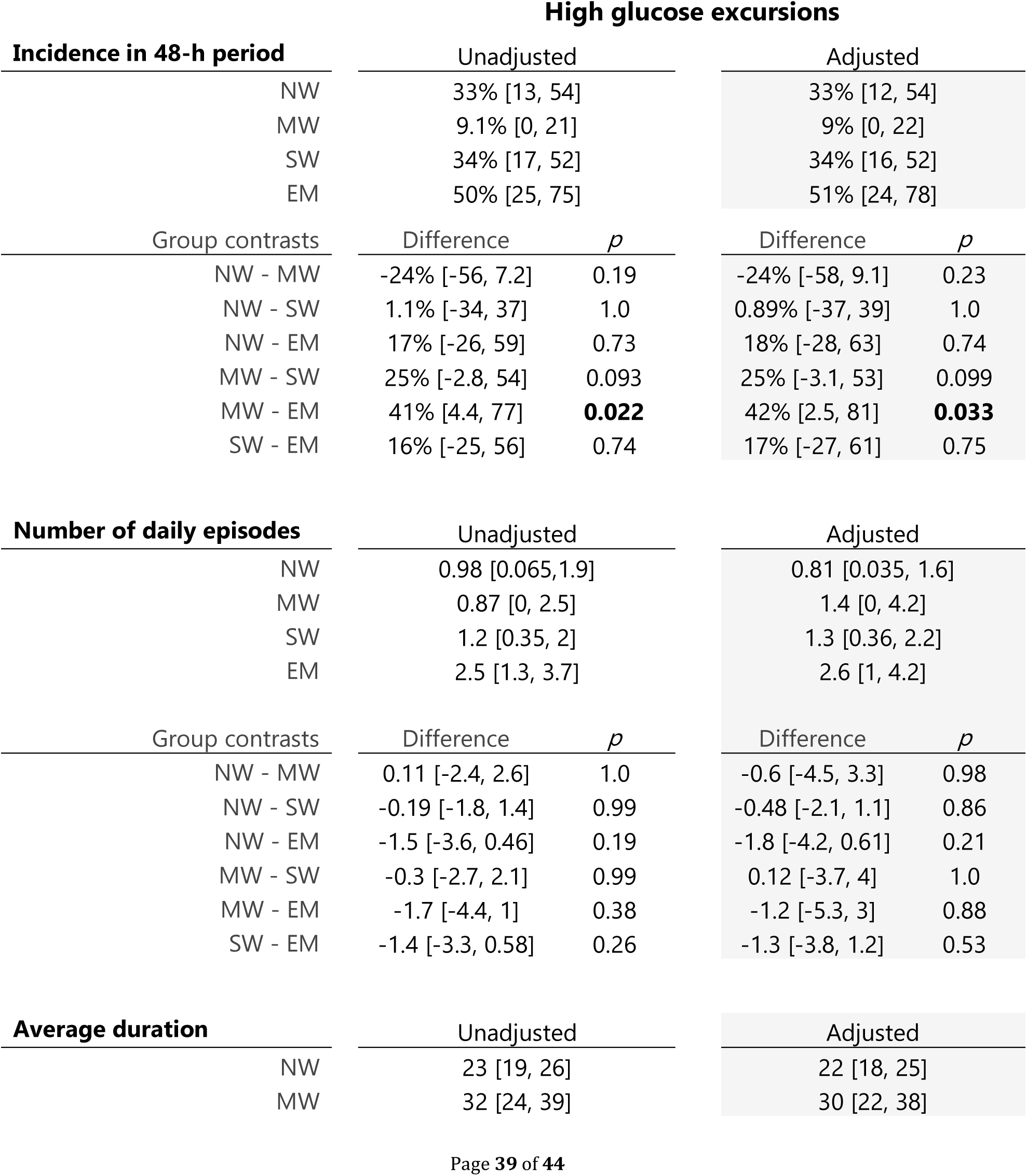
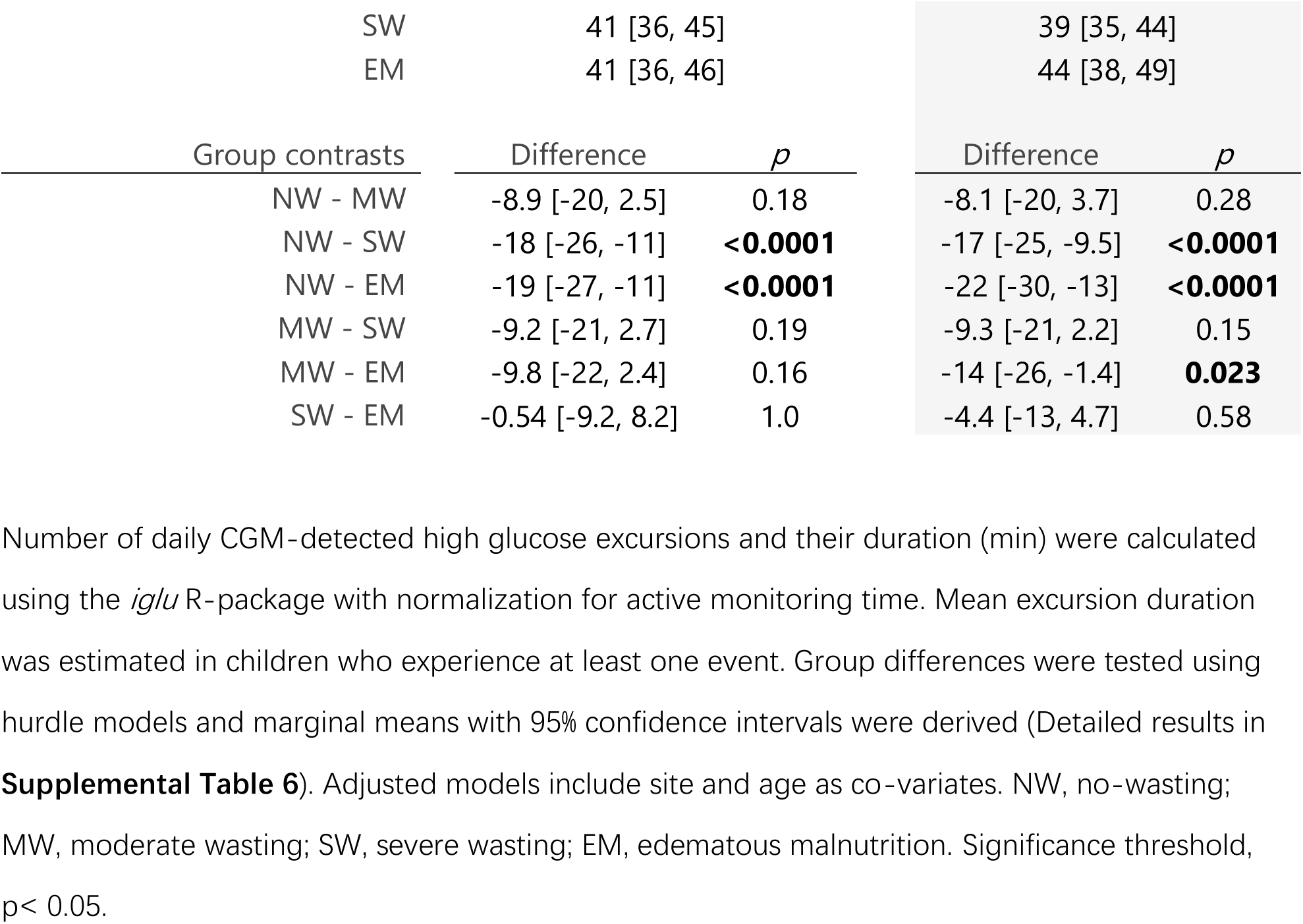
Differences in the probability, frequency, and duration of CGM-detected high glucose excursions (>11.0 mmol/L) during the 48-hour monitoring window by anthropometric status.

### High glucose variability across anthropometric groups in acutely ill children

Glucose variability was associated with anthropometric status, children with EM exhibited greater glucose instability (**Figure 3**). Individual glucose profiles, stratified by anthropometric group, are shown in Figure 3A. The median glucose levels did not differ across groups (NW, 5.6 mmol/L [6.1, 5.1]; MW, 5.2 [5.7, 4.7]; SW, 5.7 [6.2, 5.3]; EM, 6.2 [6.8, 5.6]), but children with EM exhibited significantly greater glycemic fluctuation (**Figure 3B and Table 5)**. Results were unchanged by site and age adjustment, full results presented in **Supplemental Table 8**.

**Figure 3.**
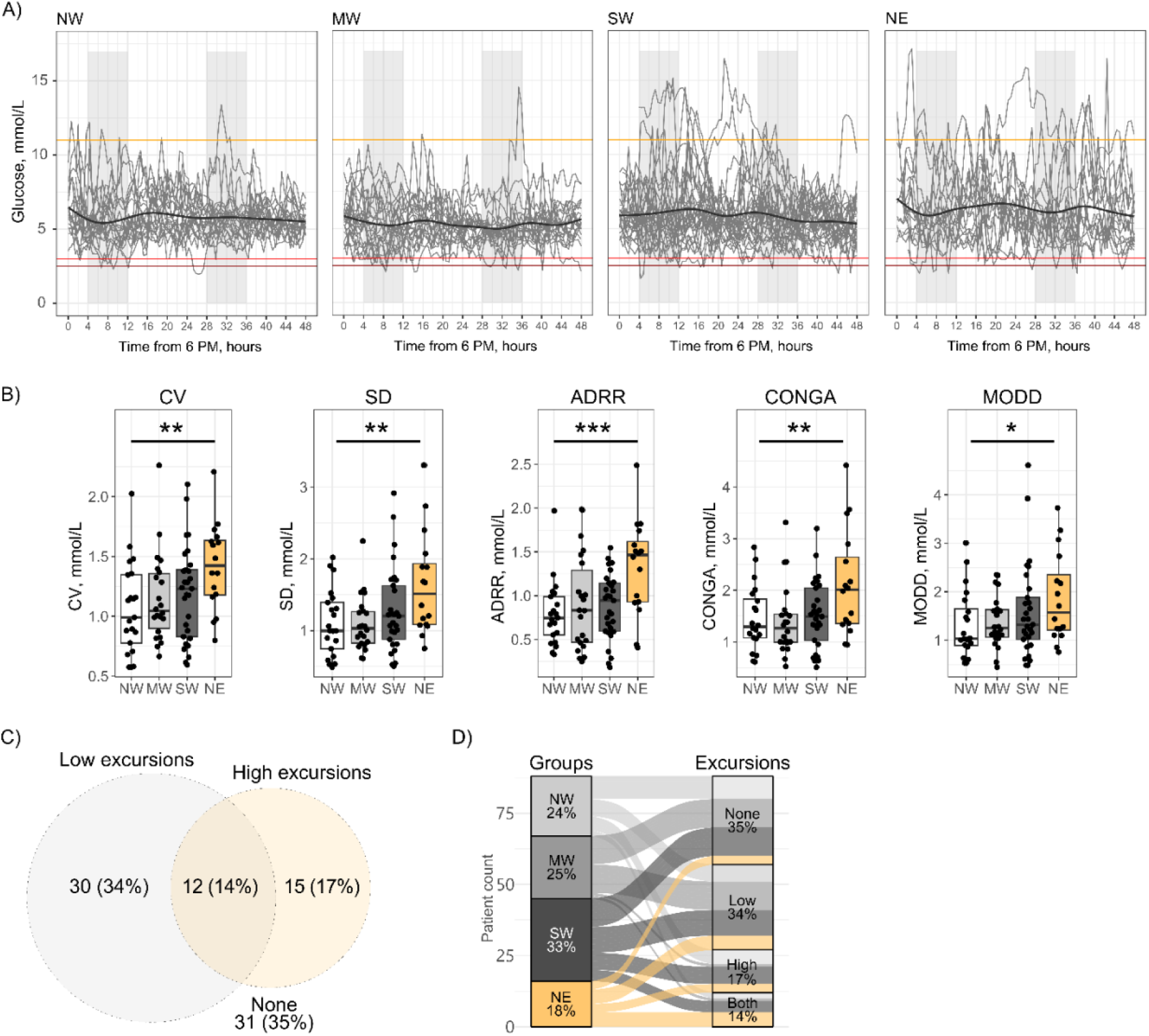
Glucose variability and glycemic patterns indices by anthropometric status. **A)** Profiles of continuous glucose monitoring (CGM) in children classified as NW, MW, SW, or EM. The black center line represents the locally estimated scatterplot smoothing (LOESS) fit for each group. Horizontal reference lines indicate thresholds for low glucose (<3.0 mmol/L; red), severely low glucose (<2.5 mmol/L; dark red), and high glucose excursions (>11.0 mmol/L; orange). Shaded grey areas denote nighttime hours (22:00 – 06:00) and X-axis represents time since 18:00 on the day of admission. **B)** Boxplots present the median of five glucose variability metrics: standard deviation (SD), coefficient of variation (CV), average daily risk range (ADRR), continuous overall/overlapping net glycemic action (CONGA) and the mean of daily differences (MODD). Metric of glucose variability were calculated using i*glu* R-package and normalized for active monitoring time. Group differences were tested using generalized linear models (Results with site and age adjustment in **Supplemental Table 6**). **C)** Venn diagram illustrating the overlap between glycemic patterns showing the number and proportion of children who maintain euglycemia, only experience low- or high-glucose excursions or experience both). **D)** Alluvial plots depicting the distribution of children across anthropometric groups that present with different glycemic patterns. Significance threshold, *p<0.05, **p<0.01, ***p<0.001. NW, no-wasting; MW, moderate wasting; SW, severe wasting; EM, edematous malnutrition.

**Table 5.**
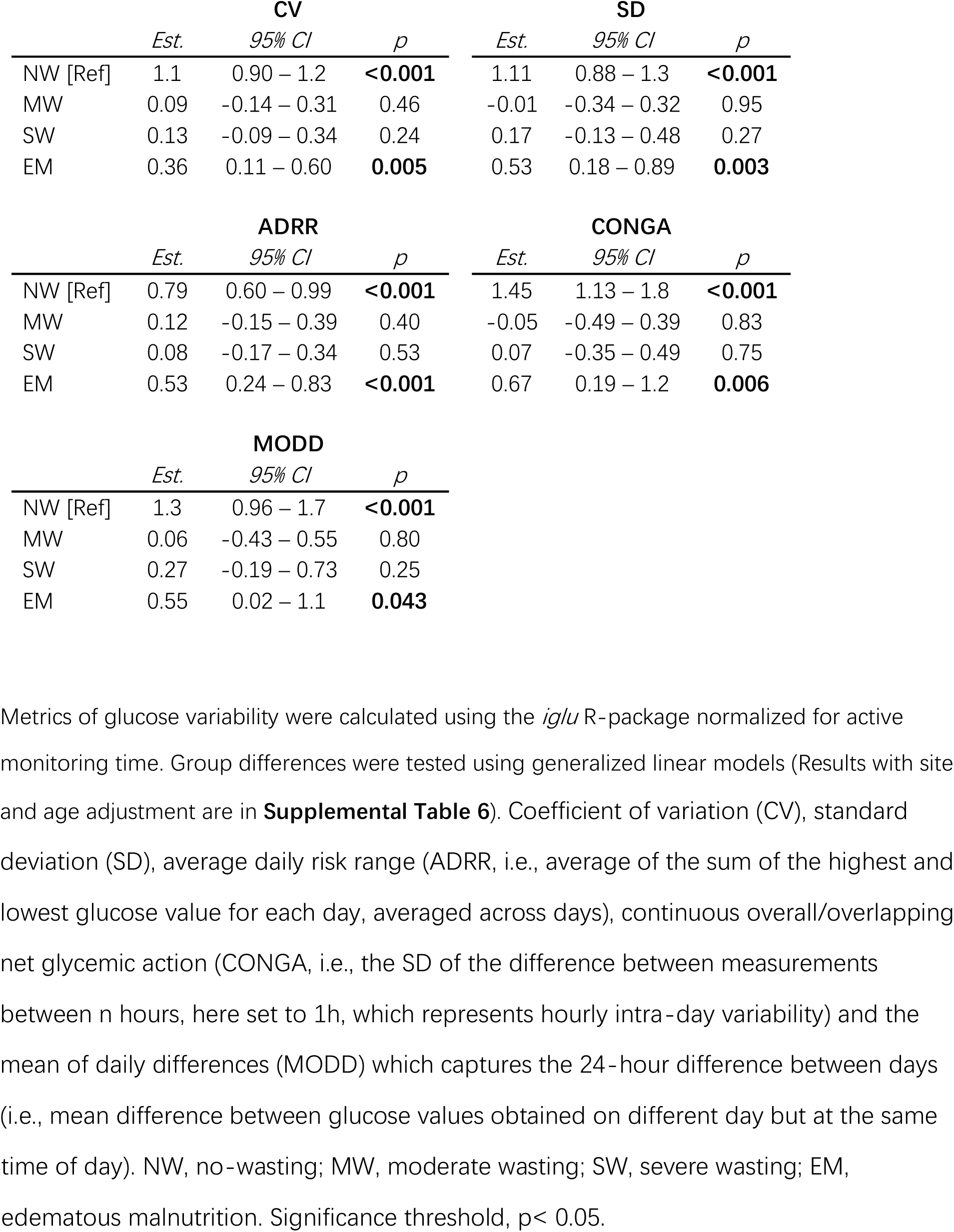
Comparison of CGM-detected glucose variability in children hospitalized with critical illness of varying anthropometric status.

|  | CV |  |  | SD |  |  |
| --- | --- | --- | --- | --- | --- | --- |
|  | *Est.* | *95% CI* | *p* | *Est.* | *95% CI* | *p* |
| NW [Ref] | 1.1 | 0.90 – 1.2 | **<0.001** | 1.11 | 0.88 – 1.3 | **<0.001** |
| MW | 0.09 | -0.14 – 0.31 | 0.46 | -0.01 | -0.34 – 0.32 | 0.95 |
| SW | 0.13 | -0.09 – 0.34 | 0.24 | 0.17 | -0.13 – 0.48 | 0.27 |
| EM | 0.36 | 0.11 – 0.60 | **0.005** | 0.53 | 0.18 – 0.89 | **0.003** |

|  | ADRR |  |  | CONGA |  |  |
| --- | --- | --- | --- | --- | --- | --- |
|  | *Est.* | *95% CI* | *p* | *Est.* | *95% CI* | *p* |
| NW [Ref] | 0.79 | 0.60 – 0.99 | **<0.001** | 1.45 | 1.13 – 1.8 | **<0.001** |
| MW | 0.12 | -0.15 – 0.39 | 0.40 | -0.05 | -0.49 – 0.39 | 0.83 |
| SW | 0.08 | -0.17 – 0.34 | 0.53 | 0.07 | -0.35 – 0.49 | 0.75 |
| EM | 0.53 | 0.24 – 0.83 | **<0.001** | 0.67 | 0.19 – 1.2 | **0.006** |

|  | MODD |  |  |
| --- | --- | --- | --- |
|  | *Est.* | *95% CI* | *p* |
| NW [Ref] | 1.3 | 0.96 – 1.7 | **<0.001** |
| MW | 0.06 | -0.43 – 0.55 | 0.80 |
| SW | 0.27 | -0.19 – 0.73 | 0.25 |
| EM | 0.55 | 0.02 – 1.1 | **0.043** |

time of day). NW, no-wasting; MW, moderate wasting; SW, severe wasting; EM, edematous malnutrition. Significance threshold, $p < 0.05$ .

Overall, 31 children (35%) maintained CGM profiles within the euglycemic range throughout the monitoring period. In contrast, 30 children (34%) exhibited only low glucose excursions, 15 (17%) only high glucose excursions, and 12 (14%) both (**Figure 3C**; **Supplemental Figure 3**; **Supplemental Table 9**). Children with euglycemic profiles had a median glucose concentration of 5.6 mmol/L (95% CI: 5.2, 5.9) and relatively low variability (IQR: 1.3 mmol/L [95% CI: 1.0, 1.5]). Children with only low glucose excursions had lower median glucose concentrations (4.9 mmol/L [95% CI: 4.5, 5.2]) but similar variability, whereas children with high glucose excursions exhibited both higher median glucose and greater variability, reflected by wider interquartile ranges and higher variability metrics (**Supplemental Table 9**).

Multinomial analysis suggested that these glycemic patterns were not related to age or site but may be associated with anthropometric status (**Figure 3D**). Children with NW tended to have lower odds of experiencing both low- and high-glucose excursions (OR: 0.25, 95% CI: 0.05, 1.2, p=0.084), while children with edema tended to have greater odds (OR: 6.7, 95% CI: 0.78, 57, p=0.082). Other clinical factors or treatments may contribute to these distinct glucose profiles, but no clear associations were identified, likely due to heterogeneity and limited sample size.

## Discussion

To our knowledge, this study is the first to use CGM to characterize patterns of dysglycemia in critically ill children across a range of anthropometric statuses. We observed frequent glucose instability early during hospitalization, suggesting that dysregulated glucose homeostasis may be an underrecognized metabolic stressor in acutely ill children in low-resource settings. Children with EM appeared to be more vulnerable, exhibiting longer low- and high-glucose excursions and greater overall variability. This study also demonstrates that CGM can be feasibly implemented in low-and middle-income country (LMIC) hospital settings.

Consistent with previous studies,^58^ CGM showed minimal systematic bias at the mean level but more limited precision at the individual-measurement level, reflected by relatively wide limits of agreement with capillary blood readings (±1.3–1.6 mmol/L), a level of discordance expected in acutely ill pediatric populations.^59^ Nearly half (47%) of children experienced at least one CGM-detected low glucose excursion early during admission, a proportion substantially higher than the 4%-14% range reported by studies using point prevalence testing in LMICs^16,27–29^ and also higher than the 19% reported in children admitted to pediatric intensive care units in the USA.^60^ However, direct comparisons with high-income settings remain challenging given differences in illness severity, and treatment intensity, particularly regarding insulin use.^58,61^ In a Belgian clinical trial, severe hypoglycemia (<2.2 mmol/L) occurred in 25% of children randomized to tight glucose control vs 1% in those without.^61^ Concerns arising from such iatrogenic hypoglycemia have reduced strict glucose targets in pediatric critical care because of potential impact on neurodevelopment.^58,61^ In our cohort, CGM-detected excursions (<2.5 mmol/L) were observed in 50% of children with EM. While CGM cannot establish clinical hypoglycemia, this finding is notable given that this threshold is associated with neurodevelopmental risk in neonates^62,63^ and that children with EM have poorer neurodevelopmental outcomes compared to those with SW.^64^

CGM-detected high glucose excursions were observed in approximately one-third of children overall and in nearly half of those with EM. While these proportions exceed point-prevalence estimates of hyperglycemia, 2-14% reported in LMIC hospital settings,^33,65,66^ our findings are comparable to the 35% reported in non-diabetic children admitted to pediatric intensive care units in the United States.^60^ Stress hyperglycemia arises from increased hepatic gluconeogenesis and insulin resistance.^67,68^ Hyperglycemia is also linked to oxidative stress and cellular damage via mitochondrial superoxide production^69^ and these effects may be amplified in severe malnutrition, a state characterized by mitochondrial dysfunction and reduced antioxidant capacity.^2,70–73^

Many studies from high resource settings have found that CGM metrics of glycemic variability are stronger predictors of poor outcome than either hyper- or hypoglycemia in pediatric intensive care patients^60,74–77^ and that variability is associated with neurodevelopmental impairment^78^ and mortality.^75–77,79,80^ Both the upward postprandial and downward inter-prandial glucose trends increase protein glycation, and oxidative stress.^74^ In cell and animal models, glycemic variability also induces inflammation, and micro- and macrovascular injury.^69,81^ which can compromise cell function inducing a vicious cycle.^81^

Glucose regulation is profoundly disrupted in severe malnutrition, owing to reduced intestinal glucose absorbtion,^18^ impaired hepatic gluconeogenesis^82^, delayed insulin secretion, and reduced glucose clearance.^83–85^ Imbalanced counter-regulations could further exacerbate glucose instability, as pancreatic inflammation and atrophy are prominent features of malnutrition.^22,82,86–89^ Thus, some children may exhibit functional insulin deficiency due to organ damage or be undiagnosed diabetics, which is not uncommon in LMICs.^90,91^ In this context, the prolonged durations of low glucose excursions observed in our cohort are comparable to those reported by CGM studies in pediatric type 1 diabetes, which document sustained low glucose excursions with median durations of approximately 30–35 minutes^92–94^ and that prolonged events exceeding two hours occur.^95^

Glucose regulation is profoundly disrupted in severe malnutrition, owing to reduced intestinal glucose absorption,¹⁸ impaired hepatic gluconeogenesis,⁸² delayed insulin secretion, and diminished peripheral glucose clearance.⁸³–⁸⁵ Dysregulated counter-regulatory responses may further exacerbate glycaemic instability, as pancreatic inflammation and atrophy are prominent features of malnutrition.²²,⁸²,⁸⁶–⁸⁹ Consequently, some children may develop functional insulin deficiency due to organ injury or have previously undiagnosed diabetes, which is not uncommon in low- and middle-income countries.⁹⁰,⁹¹ In this context, the prolonged low excursions observed in our cohort are comparable to those reported by CGM studies in pediatric type 1 diabetes, which document sustained low-glucose episodes with median durations of approximately 30–35 minutes,⁹²–⁹⁴ and in some cases exceeding two hours.⁹⁵

In this prospective observational study, we used CGM to investigate glucose regulation in an understudied pediatric population. However, several limitations should be noted. The convenience sample size was modest considering the number of comparisons across groups and sites and age, and anthropometric distribution differed between sites.

Therapeutic feeding and underlying diagnosis also varied between sites limiting condition specific analysis. Important contributors to glycemia such as medications, intravenous fluids, enteral nutrition, and feeding patterns, were not consistently recorded and biomarkers of longer-term glycemic control, hormonal regulation, oxidative stress, or inflammation were not measured. Also, while CGM alerts prompted capillary blood testing, the monitors were often held by caregivers, which could delay clinical response, which were not systematically documented. CGM devices were not specifically validated for malnourished children, sensor placement was challenging in children with severe wasting and dehydration. Finally, capillary blood glucose measurements using POC glucometers can be less accurate than lab-based arterial blood glucose measurements.^96^

## Conclusion

While some children maintain euglycemia, dysglycemic patterns were common among ill children hospitalized in LMICs. Reliance on single glucose measurements at admission likely underestimates risk and may inadequately inform clinical monitoring and management. WHO guidelines recommend early screening for hypoglycemia and pro-active management of children with severe malnutrition, including frequent feeding with F75 and provision of oral glucose if hypoglycemia is suspected.^1,24^ In practice, screening is often limited to admission or triggered by overt clinical signs, which may be insufficient to capture evolving risk in vulnerable children such as those with EM. This study provides a rationale that glucose monitoring should extend beyond admission which could prompt earlier intervention and help prevent known lasting effects of severe hypoglycemia.

Evaluating fasting from postprandial low-glucose excursions may also help to tailor feeding strategies, such as adjusting feeding intervals or the glycemic index of feeds. CGM may represent a promising tool to support optimization of therapeutic feeding formulations and improve glycemic control in critically ill children in LMIC settings.

## Data Availability

All data produced in the present work are contained in the manuscript and supplementary materials.

## Acknowledgement

We extend our sincere gratitude to all patients and families that have made this research possible. We are deeply thankful to the nurses and clinical staff working at the pediatric critical care departments at QUECH hospital in Blantyre, Malawi, and at the icddr,b in Dhaka. Without your contributions, this study would not have been possible. We would also like to acknowledge the generous funding support received by the Bill & Melinda Gates Foundation (OPP1131320). JAB is also supported by the Medical Research Council–Department for International Development–Wellcome Trust Joint Global Health Trials scheme (MR/M007367/1). Contributions from the Wellcome Trust (203077_Z_16_Z) aided in costs around staffing, facilities, and other resources. We are also grateful for our broader international research colleagues, far and wide, linked to The CHAIN Network, who have contributed directly or indirectly towards the completion of this project.

## CRediT author statement

Conceptualization ─ Ideas; formulation or evolution of overarching research goals and aims (P.P.H., C.B., F.K., K.D.T, E.B., E.S., B.K., M.B.vH, T.A., J.L.W., J.A.B., M.J.C., W.P.V, F.A., R.H.J.B.). Methodology ─ Development or design of methodology; creation of models (P.P.H., C.B., F.K., J.M.N., K.D.T, N.N., C.T., S.S.N., G.H., P.M., S.K., T.A., J.L.W., J.A.B., M.J.C., W.P.V, F.A., R.H.J.B.). Software ─ Programming, software development; designing computer programs; implementation of the computer code and supporting algorithms; testing of existing code components (P.P.H., C.B., Z.I., C.M., E.C., N.N., J.C., G.H., P.M.). Validation ─ Verification, whether as a part of the activity or separate, of the overall replication/ reproducibility of results/experiments and other research outputs (P.P.H., C.B., F.K., C.M., E.C., N.N., J.L.W., J.A.B., M.J.C., W.P.V, F.A., R.H.J.B.). Formal analysis ─ Application of statistical, mathematical, computational, or other formal techniques to analyze or synthesize study data (P.P.H., C.B., Z.I., J.C., G.H., P.M.). Investigation ─ Conducting a research and investigation process, specifically performing the experiments, or data/evidence collection (P.P.H., F.K., S.A.S., M.I., F.I., S.K., S.S.N., M.J.C., W.P.V, F.A., R.H.J.B.). Resources ─ Provision of study materials, reagents, materials, patients, laboratory samples, animals, instrumentation, computing resources, or other analysis tools (K.D.T, J.L.W., J.A.B., M.J.C., W.P.V, F.A., R.H.J.B.). Data Curation ─ Management activities to annotate (produce metadata), scrub data and maintain research data (including software code, where it is necessary for interpreting the data itself) for initial use and later reuse. (P.P.H., C.B., F.K., Z.I., C.M., E.C., N.N.). Writing - Original Draft ─ Preparation, creation and/or presentation of the published work, specifically writing the initial draft (including substantive translation) (P.P.H.). Writing - Review & Editing ─ Preparation, creation and/or presentation of the published work by those from the original research group, specifically critical review, commentary or revision – including pre-or post-publication stages (all authors). Visualization ─ Preparation, creation and/or presentation of the published work, specifically visualization/ data presentation (P.P.H., C.B.). Supervision ─ Oversight and leadership responsibility for the research activity planning and execution, including mentorship external to the core team (K.D.T, C.T., E.B., E.S., B.K., M.B.vH, T.A., J.L.W., J.A.B., M.J.C., W.P.V, F.A., R.H.J.B.). Project administration ─ Management and coordination responsibility for the research activity planning and execution (P.P.H., C.B., F.K., J.M.N., N.N., C.T., S.S.N., S.K., T.A., J.L.W., J.A.B., M.J.C., W.P.V, F.A., R.H.J.B). Funding acquisition ─ Acquisition of financial support for the project leading to this publication (J.L.W., J.A.B.).

## List of abbreviation

ATP: Adenosine triphosphate
CONGA: Continuous overall net glycemic action
CI: Confidence Interval
CV: Coefficient of variation
EM: Edematous malnutrition
IQR: Interquartile range
LOESS: Locally Estimated Scatterplot Smoothing
WAZ: Height-for-age z-score
HIV: Human Immuno-deficiency Virus
IRR: Incidence rate ratio
MW: Moderate wasting
MODD: Mean of daily differences
MUAC: Mid Upper Arm Circumference
NW: No wasting
OR: Odds ratio
SW: Severe wasting
SD: standard deviation
WHO: World Health Organisation
WAZ: Weight-for-age z-score
WHZ: Weight-for-height z-score

