## Supplemental Materials for "Patterns of Dysglycemia Identified by Continuous Glucose Monitoring among Critically Ill Children in Malawi and Bangladesh"

### Continuous Glucose Monitoring Reveals High Incidence of Dysglycemia in Children Hospitalized in Malawi and Bangladesh: Insights from a Prospective Cohort Study

#### Supplementary Materials

Sites and recruitment process ........ 3

Inclusion and exclusion criteria ........ 3

Supplementary Table 1. Description of variables included in the analysis. ........ 5

Supplementary Table 2. Description of metrics derived from the continuous glucose monitoring data and comparator values from other studies. ........ 8

Supplemental Figure 1. Study flow chart. ........ 9

Supplemental Figure 2. Pattern of continuous glucose monitoring ........ 9

Supplemental Figure 3: Bland-Altman plots showing agreement between measures obtained with continuous glucose monitoring ........ 10

Supplemental Table 3. Results from hurdle models testing differences in incidence, number and duration of hypoglycemia in children with critical illness of varying nutritional status in the 48-h period after admission. ........ 10

Supplemental Table 4. Results from hurdle models testing differences in incidence, and number of episodes of severe hypoglycemia in children with critical illness of varying nutritional status in the 48-h period after admission. ........ 15

Supplemental Table 5. Results from hurdle models testing differences in incidence, and duration of hyperglycemia in children with critical illness of varying nutritional status in the 48-h period after admission. ........ 18

Supplemental Table 6: Prevalence of hypoglycemia including number and duration of episodes obtained with continuous glucose monitoring (CGM) presented split by nutritional group. ........ 22

Supplemental Table 7. Comparison of glucose variability between children hospitalized with critical illness with varying nutritional status. ........ 24

Supplemental Table 8. Comparison of metrics describing glucose patterns in children who maintain euglycemia compared to those who experience only hypoglycemia, only hyperglycemia or both. ........ 26

#### Sites and recruitment process

Infants from two hospitals at Dhaka Hospital (Bangladesh), Queen Elizabeth Central Hospital Blantyre (Malawi). Details of the site population and disease epidemiology and the hospital characteristics were previously described.[1] For the main cohort, sites were asked to recruit approximately five children per week, 2 severely wasted/kwashiorkor (SWK), 2 moderately wasted (MW) and 1 not wasted (NW) by recruiting the first eligible child admitted in each stratum beginning on a fixed day each week.

For this sub-study, families of children were approached after prioritizing recruitment for the main cohort.

Written informed consent was obtained in participants' local language.

#### Inclusion and exclusion criteria

##### Inclusion criteria:

- Age 2 to 23 months
- Admitted to hospital with a medical illness
- Meeting criteria for one of three strata:
    - **Not wasted (NW)**: MUAC ≥12·5cm (age ≥6mo) or MUAC ≥12cm (age <6mo).
    - **Moderately wasted (MW)**: MUAC 11·5 to <12·5cm (age ≥6mo) or MUAC 11 to <12cm (age <6mo).
    - **Severely wasted or kwashiorkor (SWK)**: MUAC <11·5cm (age ≥6mo) or MUAC<11cm (age <6mo) or bilateral pedal oedema unexplained by other medical causes.

##### Exclusion criteria:

- Currently undergoing CPR or imminent cardiac arrest.
- Hospitalization for trauma
- Hospitalization for a condition requiring surgery within 6 months
- Known terminal illness expected to result in death within 6 months
- Suspected chromosomal abnormality
- Unable to tolerate oral feeds prior to current illness

- Previous inclusion of this child or a sibling in this study or any other study including the main CHAIN cohort.
- Lack of willingness to participate in follow-up visits for 6 months
- Lack of caregiver informed consent

**Supplementary Table 1.** Classification criteria of children hospitalized with acute illness.

| **Nutritional groups** | **Criteria** |
|---|---|
| No wasting (NW) | WLZ ≥-2 or,<br>if age ≥6 months, MUAC ≥12·5 cm |
| Moderately wasted (MW) | WLZ <-2 but ≥-3 or,<br>if age ≥6 months, MUAC ≥11·5 to <12·5 cm |
| Severely wasted (SW) | WLZ <-3 or,<br>if age ≥6 months, MUAC <11·5 cm |
| Edematous malnutrition (EM)[¥] | having bilateral pitting nutritional edema either: +, in feet only; ++, in feet and lower limbs, or +++, generalised including upper body and face. |

| **Other classifications** | **Criteria** |
|---|---|
| Stunted | LAZ < -2 z-score |
| Severely stunted | LAZ < -3 z-score |

Growth metrics: LAZ, length-for-age z-score; MUAC, mid-upper arm circumference; WLZ, weight-for-length z-score.

**Supplementary Table 2. Description of clinical variables.**

| Variable | Type | Unit / Categories | Description |
|---|---|---|---|
| **General Characteristics** | | | |
| Site | Categorical | Bangladesh, Malawi | |
| Sex | Categorical | Male, Female | |
| Age | Continuous | Months | calculated from difference between date of birth and date of admission |
| Prior hospitalisation | Categorical | Yes, No | as reported by primary caregiver |
| **Anthropometry** | | | |
| MUAC | Continuous | cm | Mid upper arm circumference (MUAC) |
| WLZ | Continuous | z-score | Weight-for-length z-scores (WLZ) derived for each patient with the R package 'anthro' using the 2006 WHO growth standards. Calculated separately for children with edema. |
| WAZ | Continuous | z-score | Weight-for-age z-scores (WAZ) were derived for each patient with the R package 'anthro' using the 2006 WHO growth standards. Calculated separately for children with edema. |
| LAZ | Continuous | z-score | Length-for-age z-scores (LAZ) were derived for each patient with the R package 'anthro' using the 2006 WHO growth standards |
| Stunted | Categorical | None, moderate, severe | Classification of children based on LAZ, as having: None (⩾ -2 SD), Moderate (< -2 SD) or Severe stunting (< -3 SD). |
| **Complaints on admission** | | | |
| Fever | Categorical | Yes, No | as reported by primary caregiver |
| Vomiting | Categorical | Yes, No | as reported by primary caregiver |
| Lethargy | Categorical | Yes, No | as reported by primary caregiver |
| Difficulty breathing | Categorical | Yes, No | as reported by primary caregiver |
| Diarrhea | Categorical | Yes, No | as reported by primary caregiver |
| Poor Feeding | Categorical | Yes, No | as reported by primary caregiver |

**Supplementary Table 2 (Continued).** Description of variables included in the analysis.

| Variable | Type | Unit / Categories | Description |
|---|---|---|---|
| **Clinical characteristics** | | | |
| Severe pneumonia | Categorical | Yes, No | having cough or difficulty breathing with oxygen saturation <90%, central cyanosis, or grunting; very severe chest indrawing or inability to breastfed or drink; or lethargy, reduced level of consciousness, or convulsions |
| Upper respiratory tract infection | Categorical | Yes, No | clinical observation |
| Bronchiolitis | Categorical | Yes, No | clinical observation |
| Anaemia | Categorical | None,<br>Mild,<br>Moderate/Severe | None, hemoglobin >110 g/L;<br>Mild 100–110 g/L;<br>Moderate/severe, <100 g/L |
| Acute diarrhea | Categorical | Yes, No | observed profuse watery diarrhea (i.e., 3 or more loose or watery stools in 24 hours) |
| Dehydration | Categorical | None, some, severe | Dehydration score was classified based on clinical observation of skin pinch, sunken eyes, level of consciousness, and difficulty breathing:<br>None, score of 0: without sunken eyes and no difficulty breathing and not having slow skin pinch;<br>Some, score of 1: having either sunken eyes or difficulty breathing or a slow skin pinch;<br>Severe, score of ⩾ 2: Score 2, having sunken eyes and having difficulty breathing or having sunken eyes and slow skin pinch or having difficulty breathing and a slow skin pinch; Score 3, having sunken eyes and difficulty breathing and a slow skin pinch; Score 4, having sunken eyes and difficulty breathing and a slow skin pinch and not being 'Alert'' on the AVPU scale. |
| Sepsis | Categorical | Yes, No | |
| SIRS (score >2) | Categorical | Yes, No | |
| Febrile convulsions | Categorical | Yes, No | |
| Malaria | Categorical | Yes, No | status determined by RDT test at admission |

| | | | |
|---|---|---|---|
| HIV status | Categorical | Negative, Negative-exposed, Positive | HIV status by RDT test, and status confirmed by PCR. |
| Blood glucose (mmol/L) | Ordered categorical | *Glycemia categorisation (5)* | *measures by point-of-care device at admission: Severe hypoglycemia (<3); Mild (≥3 and <3.9* mmol/L*); Normal (≥3.9 and <7* mmol/L*); Mild hyperglycemia (≥7 and <11* mmol/L*); Severe hyperglycemia (≥11* mmol/L*)* |

**Supplementary Table 3. Description of metrics derived from the continuous glucose monitoring.**

| Measures of glycemic variability | Description | Reference |
|---|---|---|
| Coefficient of Variation across all measures (CV) | indicator of daily variation (24 hour) | Rodbard (2009) |
| Standard deviation across all measures (SD) | indicator of daily variation (24 hour) | Rodbard (2009) |
| Average daily risk range (ADRR) | the average sum of the highest glucose value and the lowest glucose value for each day, with these sums averaged across days. | Kovatchev et al. (2006) |
| Continuous Overall Net Glycemic Action (CONGA) | computes the standard deviation of the difference between measurements separated by n hours, which represents hourly intra-day variability | McDonnell et al. (2005) |
| mean of daily differences (MODD) | mean difference between glucose values obtained at the same time of day | Service & Nelson (1980) |

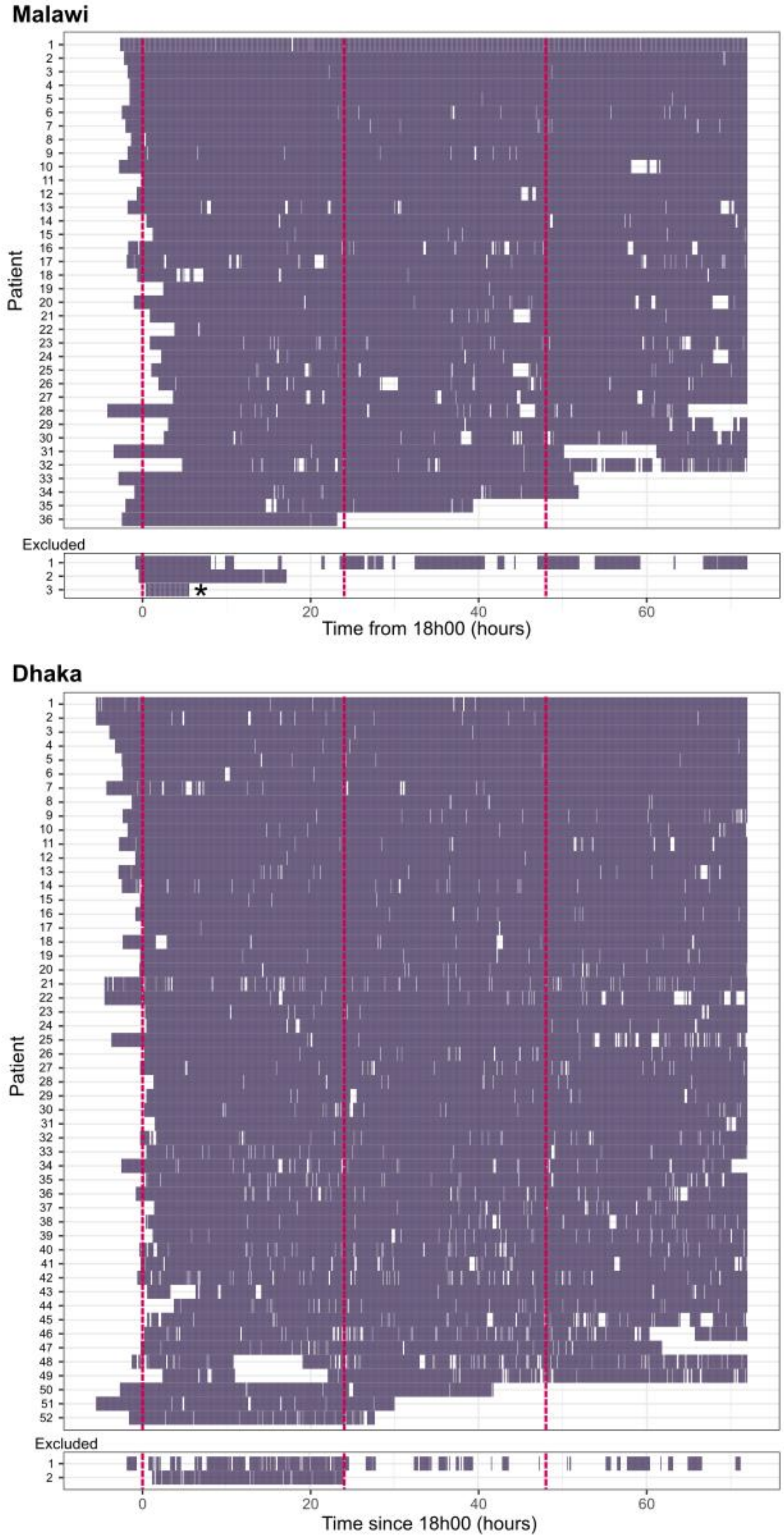


**Supplemental Figure 1. Pattern of continuous glucose monitoring** (CGM) for each participant per site (Malawi, n=36; Dhaka, n=52). Glucose profiles were excluded if less than 24 hours (n=3) or if having less than 60% coverage (n=2). Solid blocks (purple) indicate recorded data points, white spaces indicate no data recorded. Red dashed lines represent the 0, 24, and 48 hour mark from 18h00 on the day hospital admission. *****, indicates the case of early inpatient death.

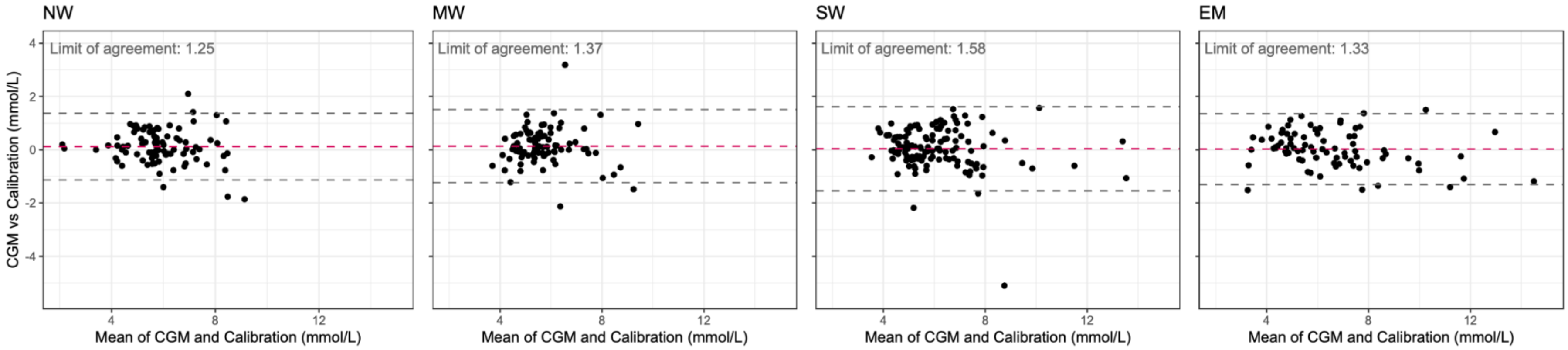


**Supplemental Figure 2: Bland-Altman plots illustrating agreement between glucose levels measured by continuous glucose monitoring (CGM) and point of care glucometer** (Calibration). The few outliers were investigated and were attributed to data entry errors from the glucometer readings. Abbreviations: NW, no-wasting; MW, moderate wasting; SW, severe wasting; EM, edematous malnutrition.

**Supplemental Table 4. Results from hurdle models testing differences in incidence, frequency and duration of CGM-detected low glucose excursions (<3 mmol/L) in critically ill children across anthropometric groups during the 48 hours of monitoring.**

| | Unadjusted | | | Adjusted | | |
|---|---|---|---|---|---|---|
| **Low glucose excursion (<3.0 mmol/L) in 48 hr window, yes/no** | *IRR* | *CI* | *p* | *IRR* | *CI* | *p* |
| No wasting [Ref] | 0.62 | 0.26 – 1.48 | 0.28 | 0.32 | 0.09 – 1.18 | 0.087 |
| Moderate wasting | 1.62 | 0.48 – 5.47 | 0.43 | 3.03 | 0.73 – 12.57 | 0.13 |
| Severe wasting | 1.15 | 0.36 – 3.62 | 0.82 | 1.79 | 0.50 – 6.41 | 0.37 |
| Edematous malnutrition | 2.71 | 0.71 – 10.36 | 0.15 | 2.98 | 0.69 – 12.75 | 0.14 |
| Malawi | | | | 2.97 | 0.96 – 9.14 | 0.058 |
| Age, months | | | | 0.99 | 0.91 – 1.08 | 0.80 |

| **Risk of ≥1 excursions in the 48-hour window** | Unadjusted | Adjusted |
|---|---|---|
| NW | 38% [17,59] | 34% [14,54] |
| MW | 50% [29,71] | 59% [38,80] |
| SW | 41% [23,60] | 47% [28,66] |
| EM | 63% [38,87] | 59% [34,84] |

| **Group contrasts** | Difference | *p* | Difference | *p* |
|---|---|---|---|---|
| NW - MW | 12% [-28,51] | 0.86 | 25% [-14,65] | 0.34 |
| NW - SW | 3.3% [-33,40] | 1.0 | 13% [-24,50] | 0.79 |
| NW - EM | 24% [-18,67] | 0.43 | 25% [-17,67] | 0.79 |
| MW - SW | -8.6% [-45,28] | 0.93 | -12% [-47,23] | 0.80 |
| MW - EM | 13% [-30,55] | 0.87 | -0.41% [-45,44] | 1.0 |
| SW - EM | 21% [-19,61] | 0.51 | 12% [-31,54] | 0.89 |

**Supplemental Table 4**. Continued

| **Number of low glucose excursions (<3.0 mmol/L) per day** | *IRR* | *CI* | *p* | *IRR* | *CI* | *p* |
|---|---|---|---|---|---|---|
| No wasting [Ref] | 1.06 | 0.47 – 2.42 | 0.88 | 1.22 | 0.47 – 3.13 | 0.68 |
| Moderate wasting | 3.03 | 1.25 – 7.39 | **0.014** | 4.26 | 1.55 – 11.73 | **0.005** |
| Severe wasting | 2 | 0.79 – 5.05 | 0.14 | 2.09 | 0.82 – 5.35 | 0.12 |
| Edematous malnutrition | 1.74 | 0.66 – 4.59 | 0.26 | 2.51 | 0.88 – 7.16 | 0.085 |
| Malawi | | | | 1.72 | 0.82 – 3.60 | 0.15 |
| Age, months | | | | 0.95 | 0.89 – 1.01 | 0.073 |

| **Number of excursions per 24hrs** | Unadjusted | Adjusted |
|---|---|---|
| NW | 1.1 [0.18,2] | 0.91 [0.13,1.7] |
| MW | 3.2 [2.1 ,4.4] | 3.9 [2,5.7] |
| SW | 2.1 [1.2, 3.1] | 1.9 [1,2.8] |
| EM | 1.9 [0.88, 2.8] | 2.3 [0.95,3.6] |

| **Group contrasts (absolute difference in number of excursions)** | Difference | *p* | Difference | *p* |
|---|---|---|---|---|
| NW - MW | -2.2 [-4.1,-0.27] | **0.019** | -3 [-5.7,-0.22] | **0.029** |
| NW - SW | -1.1 [-2.8,0.63] | 0.36 | -0.99 [-2.5,0.53] | 0.32 |
| NW - EM | -0.79 [-2.5,0.94] | 0.63 | -1.4 [-3.5,0.72] | 0.32 |
| MW - SW | 1.1 [-0.84,3] | 0.45 | 2 [-0.75,4.7] | 0.24 |
| MW - EM | 1.4 [-0.6,3.3] | 0.27 | 1.6 [-1.3,4.5] | 0.46 |
| SW - EM | 0.27 [-1.5,2] | 0.98 | -0.38 [-2.6,1.9] | 0.97 |

**Supplemental Table 4**. Continued

| **Duration of low glucose excursions (<3.0 mmol/L)** | *IRR* | *CI* | *p* | *IRR* | *CI* | *p* |
|---|---|---|---|---|---|---|
| No wasting [Ref] | 23.75 | 20.60 – 27.38 | **<0.001** | 14.82 | 12.25 – 17.93 | **<0.001** |
| Moderate wasting | 1.09 | 0.90 – 1.31 | 0.37 | 1.28 | 1.03 – 1.58 | **0.025** |
| Severe wasting | 1.01 | 0.84 – 1.21 | 0.91 | 1.17 | 0.97 – 1.41 | 0.11 |
| Edematous malnutrition | 1.59 | 1.33 – 1.89 | **<0.001** | 1.36 | 1.12 – 1.66 | **0.002** |
| Malawi | | | | 1.35 | 1.12 – 1.62 | **0.002** |
| Age, months | | | | 1.02 | 1.01 – 1.04 | **0.002** |

| **Duration** | Unadjusted | Adjusted |
|---|---|---|
| NW | 24 [20,27] | 22 [19,25] |
| MW | 26 [23,29] | 28 [24,32] |
| SW | 24 [21,27] | 26 [23,29] |
| EM | 38 [34,42] | 30 [26,34] |

| **Group contrasts (Absolute difference in minutes)** | Difference | *p* | Difference | *p* |
|---|---|---|---|---|
| NW - MW | -2.1 [-8.1,4] | 0.81 | -6 [-13,1] | 0.12 |
| NW - SW | -0.25 [-6.1,5.6] | 1.0 | -3.7 [-9.6,2.2] | 0.37 |
| NW - EM | -14 [-21,-6.9] | **<0.0001** | -7.9 [-15,-1.1] | **0.017** |
| MW - SW | 1.8 [-3.7,7.3] | 0.82 | 2.4 [-4.1,8.9] | 0.78 |
| MW - EM | -12 [-19,-5.1] | **<0.0001** | -1.9 [-8.8,5] | 0.89 |
| SW - EM | -14 [-20,-7.1] | **<0.0001** | -4.2 [-11,2.6] | 0.37 |

Hurdle models were used to separately model the occurrence (yes/no) and burden (frequency and total duration) of CGM-detected low-glucose excursions (<3 mmol/L). Marginal means with 95% confidence intervals were derived. All models account for differences in active CGM monitoring time. Adjusted models include site and age in months as co-variates. Average duration of CGM-detected low glucose excursions was calculated in children that experience at least one excursion. IRRs with 95% confidence intervals are shown; the reference-group IRR reflects the model intercept. NW, no-wasting; MW, moderate wasting; SW, severe wasting; EM, edematous malnutrition.

**Supplemental Table 5. Results from hurdle models testing differences in incidence, and frequency of CGM-detected severely low glucose excursions (<2.5 mmol/L) in critically ill children across anthropometric groups during the 48 hours of monitoring.**

| | Unadjusted | | | Adjusted | | |
|---|---|---|---|---|---|---|
| **Severely low glucose excursion (<2.5 mmol/L) in 48 hr window, yes/no** | *IRR* | *CI* | *p* | *IRR* | *CI* | *p* |
| No wasting [Ref] | 0.17 | 0.05 – 0.57 | **0.004** | 0.08 | 0.01 – 0.41 | **0.003** |
| Moderate wasting | 2.8 | 0.61 – 12.75 | 0.18 | 5.4 | 0.88 – 33.18 | 0.069 |
| Severe wasting | 0.69 | 0.13 – 3.83 | 0.67 | 1.05 | 0.17 – 6.30 | 0.96 |
| Edematous malnutrition | 6.0 | 1.25 – 28.74 | **0.025** | 6.5 | 1.14 – 36.89 | **0.035** |
| Malawi | | | | 3.01 | 0.69 – 13.05 | 0.14 |
| Age, months | | | | 1.0 | 0.89 – 1.11 | 0.94 |

| **Risk of ≥1 excursions in the 48-hour window** | Unadjusted | Adjusted |
|---|---|---|
| NW | 14% [0,29] | 12% [0,26] |
| MW | 32% [12,52] | 41% [17,66] |
| SW | 10% [0,22] | 13% [0,27] |
| EM | 50% [25,75] | 45% [20,71] |

| **Group contrasts** | Difference | *p* | Difference | *p* |
|---|---|---|---|---|
| NW - MW | 18% [-15,50] | 0.50 | 29% [-9.2,67] | 0.20 |
| NW - SW | -3.9% [-29,21] | 0.98 | 0.49% [-25,26] | 1.0 |
| NW - EM | 36% [-2.7,74] | 0.08 | 33% [-5.7,72] | 0.12 |
| MW - SW | 21% [-51,8.5] | 0.25 | -28% [-64,6.9] | 0.16 |
| MW - EM | 18% [-24,60] | 0.67 | 4.2% [-43,51] | 1.0 |
| SW - EM | 40% [3.7,76] | **0.025** | 33% [-6.9,72] | 0.14 |

**Supplemental Table 5**. Continued

| **Number of severely low glucose excursions (<2.5 mmol/L) per day** | *IRR* | *CI* | *p* | *IRR* | *CI* | *p* |
|---|---|---|---|---|---|---|
| No wasting [Ref] | 1.13 | 0.31 – 4.10 | 0.86 | 0.21 | 0.02 – 1.80 | 0.16 |
| Moderate wasting | 0.87 | 0.18 – 4.27 | 0.87 | 1.47 | 0.20 – 11.02 | 0.71 |
| Severe wasting | 1.0 | 0.16 – 6.22 | 1.0 | 1.23 | 0.16 – 9.43 | 0.84 |
| Edematous malnutrition | 0.41 | 0.06 – 2.65 | 0.35 | 0.2 | 0.01 – 2.77 | 0.23 |
| Malawi | | | | 3.64 | 0.31 – 42.15 | 0.30 |
| Age, months | | | | 1.06 | 0.86 – 1.30 | 0.60 |

| **Number of severely low glucose excursion (<2.5 mmol/L) per day** | Unadjusted | | Adjusted | |
|---|---|---|---|---|
| NW | 1.1 [0,2.6] | | 0.9 [0,2.1] | |
| MW | 0.98 [0.065,1.9] | | 1.3 [0,3.3] | |
| SW | 1.1 [0,2.6] | | 1.1 [0,2.9] | |
| EM | 0.46 [0,1.1] | | 0.18 [0,0.61] | |
| **Group contrasts** | Difference | *p* | Difference | *p* |
| NW - MW | 0.14 [-2.2,2.4] | 1.0 | -0.42 [-3.6,2.7] | 0.98 |
| NW - SW | 0 [-2.8,2.8] | 1.0 | -0.21 [-3,2.6] | 1.0 |
| NW - EM | 0.66 [-1.5,2.8] | 0.85 | 0.72 [-0.94,2.4] | 0.67 |
| MW - SW | -0.14 [-2.4,2.2] | 1.0 | 0.21 [-4.1,4.5] | 1.0 |
| MW - EM | 0.52 [-0.95,2] | 0.79 | 1.1 [-1.2,3.5] | 0.58 |
| SW - EM | 0.66 [-1.5,2.8] | 0.85 | 0.93 [-1.7,3.6] | 0.80 |

Hurdle models were used to separately model the occurrence (yes/no) and burden (frequency and total duration) of CGM-detected severely low glucose excursions (<2.5 mmol/L). Marginal means with 95% confidence intervals were derived. All models account for differences in active CGM monitoring time. Adjusted models include site and age in months as co-variates. Average duration of CGM-detected low glucose excursions was calculated in children that experience at least one excursion. IRRs with 95% confidence intervals are shown; the reference-group IRR reflects the model intercept. NW, no-wasting; MW, moderate wasting; SW, severe wasting; EM, edematous malnutrition.

**Supplemental Table 6. Results from hurdle models testing differences in incidence, and frequency of CGM-detected high glucose excursions (>11 mmol/L) in critically ill children across anthropometric groups during the 48 hours of monitoring.**

| | Unadjusted | | | Adjusted | | |
|---|---|---|---|---|---|---|
| **Experienced high glucose excursions (>11 mmol/L) in 48hrs, yes/no** | *IRR* | *CI* | *p* | *IRR* | *CI* | *p* |
| No wasting [Ref] | 0.5 | 0.20 – 1.24 | 0.13 | 0.54 | 0.15 – 1.98 | 0.36 |
| Moderate wasting | 0.2 | 0.04 – 1.11 | 0.066 | 0.2 | 0.03 – 1.23 | 0.082 |
| Severe wasting | 1.05 | 0.32 – 3.45 | 0.93 | 1.04 | 0.29 – 3.69 | 0.95 |
| Edematous malnutrition | 2 | 0.53 – 7.60 | 0.31 | 2.08 | 0.50 – 8.71 | 0.31 |
| Malawi | | | | 0.98 | 0.31 – 3.11 | 0.97 |
| Age, months | | | | 0.99 | 0.91 – 1.09 | 0.88 |

| **Risk of ≥1 excursions in the 48-hour window** | Unadjusted | Adjusted |
|---|---|---|
| NW | 33% [13,54] | 33% [12,54] |
| MW | 9.1% [0,21] | 9% [0,22] |
| SW | 34% [17,52] | 34% [16,52] |
| EM | 50% [25,75] | 51% [24,78] |

| **Group contrasts** | Difference | *p* | Difference | *p* |
|---|---|---|---|---|
| NW - MW | -24% [-56,7.2] | 0.19 | -24% [-58,9.1] | 0.23 |
| NW - SW | 1.1% [-34,37] | 1.0 | 0.89% [-37,39] | 1.0 |
| NW - EM | 17% [-26,59] | 0.73 | 18% [-28,63] | 0.74 |
| MW - SW | 25% [-2.8,54] | 0.093 | 25% [-3.1,53] | 0.099 |
| MW - EM | 41% [4.4,77] | **0.022** | 42% [2.5,81] | **0.033** |
| SW - EM | 16% [-25,56] | 0.74 | 17% [-27,61] | 0.75 |

**Supplemental Table 6**. Continued

| **Number of high glucose excursions (>11 mmol/L) per 24hrs** | *IRR* | *CI* | *p* | *IRR* | *CI* | *p* |
|---|---|---|---|---|---|---|
| No wasting [Ref] | 0.98 | 0.39 – 2.47 | 0.97 | 0.61 | 0.19 – 1.98 | 0.41 |
| Moderate wasting | 0.89 | 0.11 – 7.01 | 0.91 | 1.73 | 0.19 – 15.90 | 0.63 |
| Severe wasting | 1.19 | 0.38 – 3.77 | 0.76 | 1.59 | 0.49 – 5.11 | 0.44 |
| Edematous malnutrition | 2.57 | 0.91 – 7.24 | 0.073 | 3.19 | 1.00 – 10.18 | **0.049** |
| Malawi | | | | 2.31 | 0.91 – 5.84 | 0.077 |
| Age, months | | | | 0.98 | 0.92 – 1.04 | 0.53 |

| **Number of excursions per 24hrs** | Unadjusted | Adjusted |
|---|---|---|
| NW | 0.98 [0.065,1.9] | 0.81 [0.035,1.6] |
| MW | 0.87 [0,2.5] | 1.4 [0,4.2] |
| SW | 1.2 [0.35,2] | 1.3 [0.36,2.2] |
| EM | 2.5 [1.3,3.7] | 2.6 [1,4.2] |

| **Group contrasts** | Difference | *p* | Difference | *p* |
|---|---|---|---|---|
| NW - MW | 0.11 [-2.4,2.6] | 1.0 | -0.6 [-4.5,3.3] | 0.98 |
| NW - SW | -0.19 [-1.8,1.4] | 0.99 | -0.48 [-2.1,1.1] | 0.86 |
| NW - EM | -1.5 [-3.6,0.46] | 0.19 | -1.8 [-4.2,0.61] | 0.21 |
| MW - SW | -0.3 [-2.7,2.1] | 0.99 | 0.12 [-3.7,4] | 1.0 |
| MW - EM | -1.7 [-4.4,1] | 0.38 | -1.2 [-5.3,3] | 0.88 |
| SW - EM | -1.4 [-3.3,0.58] | 0.26 | -1.3 [-3.8,1.2] | 0.53 |

**Supplemental Table 6**. Continued

| Duration of high glucose excursions (>11 mmol/L) | *IRR* | *CI* | *p* | *IRR* | *CI* | *p* |
|---|---|---|---|---|---|---|
| No wasting [Ref] | 22.57 | 19.31 – 26.38 | **<0.001** | 28.58 | 23.51 – 34.74 | **<0.001** |
| Moderate wasting | 1.4 | 1.04 – 1.87 | **0.025** | 1.37 | 1.00 – 1.88 | 0.051 |
| Severe wasting | 1.81 | 1.49 – 2.18 | **<0.001** | 1.8 | 1.46 – 2.21 | **<0.001** |
| Edematous malnutrition | 1.83 | 1.51 – 2.22 | **<0.001** | 2 | 1.63 – 2.46 | **<0.001** |
| Malawi | | | | 0.95 | 0.80 – 1.13 | 0.57 |
| Age, months | | | | 0.98 | 0.96 – 0.99 | **0.001** |

| **Duration** | Counts | Counts |
|---|---|---|
| NW | 23 [19,26] | 22 [18,25] |
| MW | 32 [24,39] | 30 [22,38] |
| SW | 41 [36,45] | 39 [35,44] |
| EM | 41 [36,46] | 44 [38,49] |

| **Group contrasts** | Difference | *p* | Difference | *p* |
|---|---|---|---|---|
| NW - MW | -8.9 [-20,2.5] | 0.18 | -8.1 [-20,3.7] | 0.28 |
| NW - SW | -18 [-26,-11] | **<0.0001** | -17 [-25,-9.5] | **<0.0001** |
| NW - EM | -19 [-27,-11] | **<0.0001** | -22 [-30,-13] | **<0.0001** |
| MW - SW | -9.2 [-21,2.7] | 0.19 | -9.3 [-21,2.2] | 0.15 |
| MW - EM | -9.8 [-22,2.4] | 0.16 | -14 [-26,-1.4] | **0.023** |
| SW - EM | -0.54 [-9.2,8.2] | 1.0 | -4.4 [-13,4.7] | 0.58 |

Hurdle models were used to separately model the occurrence (yes/no) and burden (frequency and total duration) of CGM-detected high-glucose excursions (>11 mmol/L). Marginal means with 95% confidence intervals were derived. All models account for differences in active CGM monitoring time. Adjusted models include site and age in months as co-variates. Average duration of CGM-detected high glucose excursions was calculated in children that experience at least one excursion. IRRs with 95% confidence intervals are shown; the reference-group IRR reflects the model intercept. NW, no-wasting; MW, moderate wasting; SW, severe wasting; EM, edematous malnutrition.

**Supplemental Table 7:** Prevalence of CGM-detected low and severely low glucose excursions (<3.0 mmol/L) and (<2.5 mmol/L) with number and duration, stratified by anthropometric group. Outcomes are summarised for the full 48-hour monitoring time and separately for day- and nighttime hours.

| | | | SAM patients | |
|---|---|---|---|---|
| **Low glucose excursions (<3.0 mmol/L)** | **NW** n = 21 | **MW** n = 22 | **SW** n = 29 | **EM** n = 16 |
| **48hrs** | 8 (38%) | 11 (50%) | 13 (45%) | 10 (63%) |
| If yes, | | | | |
| Measures | 6.5 (3.8, 19) | 34 (7.5, 47) | 11 (1.0, 30) | 19 (16, 25) |
| Number of episodes | 0.59 (0.33, 0.84) | 1.3 (0.67, 2.5) | 1.0 (0.33, 2.0) | 0.84 (0.42, 1.6) |
| Duration, min | 20 (19, 23) | 21 (16, 34) | 16 (7.5, 35) | 29 (21, 63) |
| **Day** | 5 (24%) | 6 (27%) | 11 (38%) | 9 (56%) |
| If yes, | | | | |
| Measures | 4.0 (3.0, 9.0) | 22 (11, 38) | 8.0 (5.0, 30) | 15 (12, 17) |
| Number of episodes | 0.33 (0.33, 0.50) | 1.2 (1.0, 2.1) | 1.0 (0.50, 1.0) | 0.67 (0.33, 1.3) |
| Duration, min | 20 (15, 26) | 22 (13, 30) | 18 (10, 39) | 28 (19, 38) |
| **Night** | 5 (24%) | 10 (45%) | 8 (28%) | 8 (50%) |
| If yes, | | | | |
| Measures | 8.0 (4.0, 9.0) | 22 (2.8, 29) | 4.5 (1.0, 12) | 8.5 (3.8, 14) |
| Number of episodes | 0.67 (0.33, 0.67) | 1.0 (0.67, 1.6) | 0.33 (0.33, 0.84) | 0.33 (0.33, 0.42) |
| Duration, min | 20 (20, 25) | 23 (11, 35) | 15 (5.0, 31) | 25 (18, 48) |
| **Severely low glucose excursions (<2.5 mmol/L)** | | | | |
| **48hrs** | 3 (14%) | 7 (32%) | 3 (10%) | 8 (50%) |
| If severe yes, | | | | |
| Count of measures | 12 (6.5, 22) | 7.0 (3.0, 18) | 13 (8.0, 14) | 7.5 (5.5, 9.3) |
| Number of episodes | 0.33 (0.33, 0.83) | 0.67 (0.33, 1.0) | 1.0 (0.67, 1.0) | 0.50 (0.33, 0.67) |
| Duration, min | 15 (10, 73) | 18 (10, 30) | 20 (19, 23) | 20 (17, 34) |
| **Day** | 2 (9.5%) | 3 (14%) | 3 (10%) | 7 (44%) |
| If severe yes, | | | | |
| Measures | 21 (13, 29) | 6.0 (4.5, 13) | 10 (5.5, 12) | 3.0 (1.5, 6.0) |
| Number of episodes | 0.50 (0.42, 0.59) | 0.67 (0.50, 0.67) | 0.67 (0.50, 0.84) | 0.33 (0.33, 0.33) |
| Duration, min | 71 (42, 101) | 15 (10, 26) | 23 (14, 25) | 10 (5.8, 30) |
| **Night** | 2 (9.5%) | 6 (27%) | 2 (6.9%) | 5 (31%) |
| If severe yes, | | | | |
| Measures | 4.0 (2.5, 5.5) | 7.5 (4.0, 12) | 2.5 (2.3, 2.8) | 6.0 (5.0, 6.0) |
| Number of episodes | 0.50 (0.42, 0.59) | 0.67 (0.42, 0.67) | 0.33 (0.33, 0.33) | 0.33 (0.33, 0.33) |
| Duration, min | 11 (8.1, 14) | 23 (12, 29) | 13 (11, 14) | 30 (20, 30) |

Counts are normalized to active monitoring time. Day-night cycle, 06:00 to 22:00. For the day-night comparison, the number of episodes were normalized per 8 hours to account for difference in time window and adjusted for monitoring time. Abbreviations: NW, no-wasting; MW, moderate wasting; SW, severe wasting; EM, edematous malnutrition.

**Supplemental Table 8. Glucose variability across anthropometric groups in critically ill children.**

**CV**

| | **Unadjusted** | | | **Adjusted** | | |
|---|---|---|---|---|---|---|
| | *Est.* | *CI* | *p* | *Est.* | *CI* | *p* |
| NW [Ref] | 1.06 | 0.90 – 1.23 | **<0.001** | 1.04 | 0.81 – 1.27 | **<0.001** |
| MW | 0.09 | -0.14 – 0.31 | 0.456 | 0.13 | -0.12 – 0.39 | 0.299 |
| SW | 0.13 | -0.09 – 0.34 | 0.243 | 0.16 | -0.07 – 0.39 | 0.17 |
| EM | 0.36 | 0.11 – 0.60 | **0.005** | 0.38 | 0.11 – 0.64 | **0.005** |
| Site, Malawi | | | | 0.09 | -0.11 – 0.29 | 0.394 |
| Age, months | | | | 0 | -0.02 – 0.01 | 0.636 |

**SD**

| | **Unadjusted** | | | **Adjusted** | | |
|---|---|---|---|---|---|---|
| | *Est.* | *CI* | *p* | *Est.* | *CI* | *p* |
| NW [Ref] | 1.11 | 0.88 – 1.34 | **<0.001** | 1.09 | 0.76 – 1.42 | **<0.001** |
| MW | -0.01 | -0.34 – 0.32 | 0.951 | 0.02 | -0.34 – 0.39 | 0.91 |
| SW | 0.17 | -0.13 – 0.48 | 0.266 | 0.19 | -0.13 – 0.52 | 0.244 |
| EM | 0.53 | 0.18 – 0.89 | **0.003** | 0.54 | 0.16 – 0.92 | **0.005** |
| Site, Malawi | | | | 0.06 | -0.23 – 0.35 | 0.698 |
| Age, months | | | | 0 | -0.02 – 0.02 | 0.86 |

**ADRR**

| | **Unadjusted** | | | **Adjusted** | | |
|---|---|---|---|---|---|---|
| | *Est.* | *CI* | *p* | *Est.* | *CI* | *p* |
| NW [Ref] | 0.79 | 0.60 – 0.99 | **<0.001** | 0.74 | 0.48 – 1.01 | **<0.001** |
| MW | 0.12 | -0.15 – 0.39 | 0.396 | 0.26 | -0.04 – 0.55 | 0.091 |
| SW | 0.08 | -0.17 – 0.34 | 0.532 | 0.17 | -0.09 – 0.44 | 0.199 |
| EM | 0.53 | 0.24 – 0.83 | **<0.001** | 0.6 | 0.29 – 0.90 | **<0.001** |
| Site, Malawi | | | | 0.25 | 0.02 – 0.49 | **0.036** |
| Age, months | | | | -0.01 | -0.03 – 0.01 | 0.202 |

**CONGA**

| | **Unadjusted** | | | **Adjusted** | | |
|---|---|---|---|---|---|---|
| | *Est.* | *CI* | *p* | *Est.* | *CI* | *p* |
| NW [Ref] | 1.45 | 1.13 – 1.77 | **<0.001** | 1.36 | 0.91 – 1.80 | **<0.001** |
| MW | -0.05 | -0.49 – 0.39 | 0.828 | 0.02 | -0.47 – 0.52 | 0.928 |
| SW | 0.07 | -0.35 – 0.49 | 0.752 | 0.12 | -0.32 – 0.56 | 0.596 |
| EM | 0.67 | 0.19 – 1.16 | **0.006** | 0.66 | 0.15 – 1.18 | **0.011** |
| Site, Malawi | | | | 0.14 | -0.25 – 0.53 | 0.483 |
| Age, months | | | | 0 | -0.03 – 0.03 | 0.968 |

**MODD**

| | **Unadjusted** | | | **Adjusted** | | |
|---|---|---|---|---|---|---|
| | *Est.* | *CI* | *p* | *Est.* | *CI* | *p* |
| NW [Ref] | 1.32 | 0.96 – 1.67 | **<0.001** | 1.3 | 0.81 – 1.79 | **<0.001** |
| MW | 0.06 | -0.43 – 0.55 | 0.802 | 0.14 | -0.40 – 0.68 | 0.611 |
| SW | 0.27 | -0.19 – 0.73 | 0.246 | 0.32 | -0.17 – 0.81 | 0.196 |
| EM | 0.55 | 0.02 – 1.08 | **0.043** | 0.59 | 0.02 – 1.15 | **0.041** |
| Site, Malawi | | | | 0.15 | -0.28 – 0.58 | 0.502 |
| Age, months | | | | -0.01 | -0.04 – 0.03 | 0.643 |

Metric of glucose variability were calculated using *iglu* R package, accounting for differences in active monitoring time. Group differences were tested using generalized linear models with or without adjustment for site and age. Standard deviation (SD), coefficient of variation (CV), average daily risk range (ADRR, i.e., average of the sum of the highest and lowest glucose value for each day, averaged across days), continuous overall/overlapping net glycemic action (CONGA, i.e., the SD of the difference between measurements between n hours, here set to 1h, which represents hourly intra-day variability) and the mean of daily differences (MODD) which captures the 24-hour difference between days (i.e., mean difference between glucose values obtained on different day but at the same time of day). NW, no-wasting; MW, moderate wasting; SW, severe wasting; EM, edematous malnutrition. Significance threshold, $P < 0.05$.

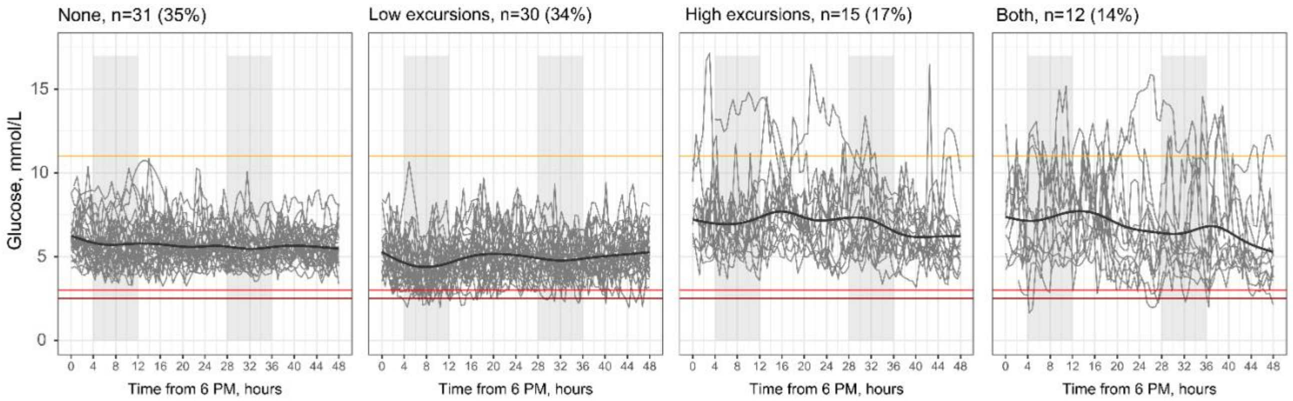


**Supplemental Figure 3**. Comparison of glucose variability patterns of continuous glucose monitoring (CGM) data measured in children who maintained CGM-readings within the euglycemic range versus those who experienced either: only low glucose excursions, only high excursions or both. The black center line presents the locally estimated scatterplot smoothing (LOESS) fit for the group. Low glucose thresholds in red (<3.0 mmol/L) and severely low in dark red (<2.5 mmol/L); high glucose threshold in orange (>11 mmol/L). Grey shadowed areas present nighttime hours between 22h:00 and 06h0 and X-axis tallies time since 18h:00 on the day of admission.

**Supplemental Table 9. Comparison of metrics describing glucose patterns in children who maintain CGM-readings in the euglycemia range compared to those who experience only low glucose excursions, only high excursions or both.**

| | Median | | | IQR | | |
|---|---|---|---|---|---|---|
| | *Est.* | *CI* | *p* | *Est.* | *CI* | *p* |
| Euglycemic | 5.57 | 5.20 – 5.94 | **<0.001** | 1.26 | 1.01 – 1.51 | **<0.001** |
| Low excursions | -0.7 | -1.23 – -0.17 | **0.009** | 0.14 | -0.22 – 0.50 | 0.446 |
| High excursions | 1.22 | 0.57 – 1.87 | **<0.001** | 0.82 | 0.38 – 1.26 | **<0.001** |
| Both | 0.91 | 0.21 – 1.61 | **0.011** | 1.47 | 1.00 – 1.95 | **<0.001** |

| | CV | | | SD | | |
|---|---|---|---|---|---|---|
| | *Est.* | *CI* | *p* | *Est.* | *CI* | *p* |
| Euglycemic | 0.89 | 0.80 – 0.98 | **<0.001** | 0.92 | 0.79 – 1.04 | **<0.001** |
| Low excursions | 0.32 | 0.19 – 0.45 | **<0.001** | 0.15 | -0.03 – 0.33 | 0.109 |
| High excursions | 0.39 | 0.23 – 0.55 | **<0.001** | 0.69 | 0.47 – 0.91 | **<0.001** |
| Both | 0.94 | 0.77 – 1.11 | **<0.001** | 1.29 | 1.05 – 1.53 | **<0.001** |

| | ADRR | | | CONGA | | |
|---|---|---|---|---|---|---|
| | *Est.* | *CI* | *p* | *Est.* | *CI* | *p* |
| Euglycemic | 0.49 | 0.39 – 0.60 | **<0.001** | 1.08 | 0.90 – 1.26 | **<0.001** |
| Low excursions | 0.71 | 0.56 – 0.86 | **<0.001** | 0.34 | 0.08 – 0.59 | **0.01** |
| High excursions | 0.39 | 0.21 – 0.58 | **<0.001** | 0.88 | 0.57 – 1.19 | **<0.001** |
| Both | 1.06 | 0.85 – 1.26 | **<0.001** | 1.74 | 1.40 – 2.08 | **<0.001** |

| | MODD | | |
|---|---|---|---|
| | *Est.* | *CI* | *p* |
| Euglycemic | 1.09 | 0.87 – 1.31 | **<0.001** |
| Low excursions | 0.22 | -0.10 – 0.54 | 0.172 |
| High excursions | 0.82 | 0.44 – 1.21 | **<0.001** |
| Both | 1.58 | 1.17 – 2.00 | **<0.001** |

Metric of glucose variability were calculated using *iglu* R package accounting for differences in active monitoring. Group differences were tested using generalized linear models. Interquartile range (IQR), standard deviation (SD), coefficient of variation (CV), average daily risk range (ADRR, i.e., average of the sum of the highest and lowest glucose value for each day, averaged across days), continuous overall/overlapping net glycemic action (CONGA, i.e., the SD of the difference between measurements between n hours, here set to 1h, which represents hourly intra-day variability) and the mean of daily differences (MODD) which captures the 24-hour difference between days (i.e., mean difference between glucose values obtained on different day but at the same time of day). Significance threshold, $P < 0.05$.
